# Frequent whole blood donations select for DNMT3A variants mediating enhanced response to erythropoietin

**DOI:** 10.1101/2022.07.24.22277825

**Authors:** D. Karpova, H. Huerga Encabo, E. Donato, I. Kotova, S. Calderazzo, AM. Leppä, J. Panten, A. Przbylla, E. Seifried, A. Kopp-Schneider, TN. Wong, D. Bonnet, H. Bonig, A. Trumpp

## Abstract

**Background:** Blood donation saves lives. Provided they are in good health, male volunteers can donate as often as six times per year from the age of 18 into their late sixties. The burden of blood donation is very unevenly distributed, with a small minority of altruistic individuals providing this critical resource. While the consequences of persistent iron depletion in blood donors have been studied in the context of cancer and coronary heart disease, potential effects of the erythropoietic stress from repetitive large-volume phlebotomy remain unexplored. We sought to investigate if and how repeated blood donations affect the clonal composition of the hematopoietic stem and progenitor cell (HSPC) compartment.

**Methods:** 105 healthy, male individuals with an extensive blood donation history (median of **120 donations** per donor; median age of 66 yrs.) were screened for the presence of clonal hematopoiesis (CH) using a sequencing panel covering 141 genes commonly mutated in human myeloid neoplasms. The control cohort consisted of 103 healthy, male donors with a median of **5 donations** per donor and a median age of 63. Donors positive for CH were subsequently studied longitudinally. The pathogenicity of detected variants was compared using established scoring systems. Finally, to assess the functional consequences of blood-donation induced CH, selected CH mutations were introduced by CRISPR-mediated editing into HSPCs from human cord blood (CB) or bone marrow (BM). The effect of these mutations was tested under different stress stimuli using functional *ex vivo* long-term culture initiating cells (LTC-IC) assays.

**Results:** Compared to the control cohort, frequent donors were significantly more likely to have mutations in genes encoding for epigenetic modifiers (44.7 vs. 22.3 %), most specifically in the two genes most commonly mutated in CH, *DNMT3A* and *TET2* (35.2 vs. 20.3 %). However, no difference in the variant allele frequency (VAF) of detected mutations was found between the groups. Longitudinal analysis revealed that the majority of the mutations remained at a stable VAF over an observation period of approximately one year. Three *DNMT3A* variants from the frequent donor cohort were introduced into healthy HSPCs and functionally analyzed: All expanded in response to EPO, but none responded to LPS or IFNγ stimulation. This contrasted with the leukemogenic *DNMT3A* R882H mutation, which did not expand in the presence of EPO but instead responded strongly to inflammatory stimuli.

**Conclusions:** Frequent whole blood donation is associated with a higher prevalence of CH driven by mutations in genes encoding for epigenetic modifiers, with *DNMT3A* and *TET2* being the most common. This increased CH prevalence is not associated with a higher pathogenicity of the associated variants and is likely a result of the selection of clones with improved responsiveness to EPO under the condition of bleeding stress. Our data show that even highly frequent blood donations over many years is not increasing the risk for malignant clones further underscoring the safety of repetitive blood donations. To our knowledge, this is the first CH study analyzing a cohort of individuals known for their superior health and survival, able to donate blood until advanced age. Thus, our analysis possibly identified mutations associated with beneficial outcomes, rather than a disease condition, such as mutations in *DNMT3A* that mediated the improved expansion of HSPCs in EPO enriched environments. Our data support the notion of ongoing Darwinian evolution in humans at the somatic stem cell level and present EPO as one of the environmental factors to which HSPCs with specific mutations may respond with superior fitness.

## Introduction

Access to large quantities of blood products is a backbone of modern Western medicine. If all eligible citizens shared the load of blood donation equally, each person would need to donate once every ten years. Currently, the burden of blood donation is shouldered by a very small group of altruistic healthy volunteers with approximately 4% of eligible donors donating, on average, 1.9 units per year^1^. In Germany, at the Red Cross Blood Donation Service Baden-Wuerttemberg-Hessen alone, approximately 1.5 million whole blood donations are collected each year. The limits of 4 (women) or 6 (men) whole blood donations per year aim to protect the iron stores of donors from critical depletion and render blood donations acutely safe^2^. Beyond iron homeostasis^3-6^, little attention has been paid to long-term donor outcomes. One unit of blood, 500 ml, represents approximately 10-15% of a donor’s total blood volume^7^. Thus, donation of one unit of blood per year increases the annual red blood cell (RBC) turnover rate by 2.5-5%, and donation of six units per year by six times this number. The long-term hematopoietic consequences of erythropoietic stress from regular blood donation have not been addressed appropriately. A recent nationwide study of Swedish blood donors reported a modest decrease of the risk of acute myeloid leukemia (AML), although overall protective effects of blood donation against disease remain poorly documented^8^.

Hematopoietic stem cells (HSCs) have been shown to be kept in reversible deep quiescence, called dormancy state, during healthy homeostasis^9-11^. Different stressors and inflammatory stimuli such as interferons (IFN), lipopolysaccharide (LPS), chemotherapy or blood loss trigger exit of HSCs from dormancy and entry into active cycling^9,12-14^. Transition from dormancy to active proliferation leads to increased replicative stress, which in turn accelerates the acquisition and propagation of genetic lesions^15^. The expansion of HSC clones (and their progeny) carrying somatic mutations is a recently characterized phenomenon referred to as clonal hematopoiesis (CH) that becomes particularly apparent during aging^16-20^. Preservation of the dormant state of HSCs during homeostasis therefore counteracts the emergence of CH, while proliferative signals from the environment promote it^15^. For example, IFNγ has been reported to favor the outgrowth of HSC clones with mutations in the epigenetic driver *DNMT3A*^21^. Furthermore, *DNMT3A* mutated HSCs have been shown to expand in the context of stem cell transplantation^22,23^, whereas HSCs carrying mutations in the DNA damage response genes PPM1D^24^ and TP53^25^ expand following chemotherapy. We hypothesized that repeated regular exposure to blood loss associated stress might impact the clonal composition of the hematopoietic stem and progenitor cell (HSPC) pool over time.

## Results

### Frequent blood donors exhibit similar characteristics of clonal hematopoiesis to control

We performed targeted next-generation sequencing of DNA isolated from peripheral blood (PB) leukocytes of >60-year-old, male blood donors who had donated frequently (Frequent Donor (FD) Cohort) throughout their adult life with a median of 120 total donations per donor (see Table 1 and Supplemental Table 1 for detailed characteristics of the cohort). The control cohort was comprised of donors of similar age with very few lifetime donations (Control Donor (CD) Cohort; median of 5 total donations per donor; see Figure 1/Table 1 and Supplemental Table 1). Male donors were chosen for the study due to a higher limit of maximum donations per year (maximum of 6 vs. 4 units per year for male vs. female donors, respectively^2^) and therefore, on average, potential for a higher exposure to erythropoietic stress. Moreover, the variability in female menstrual patterns would have complicated the analysis in females. The median donor age at the time of the index donation was 66 years vs. 63 years in the FD vs. CD cohort, respectively. Distinct from most CH studies published to date^16,17,26-32^, both our cohorts consisted exclusively of closely monitored, exceptionally healthy individuals. A broad unique molecular identifier (UMI)-based sequencing panel, covering 141 genes known to be mutated in hematologic malignancies, was chosen for targeted sequencing.

**Figure 1.**
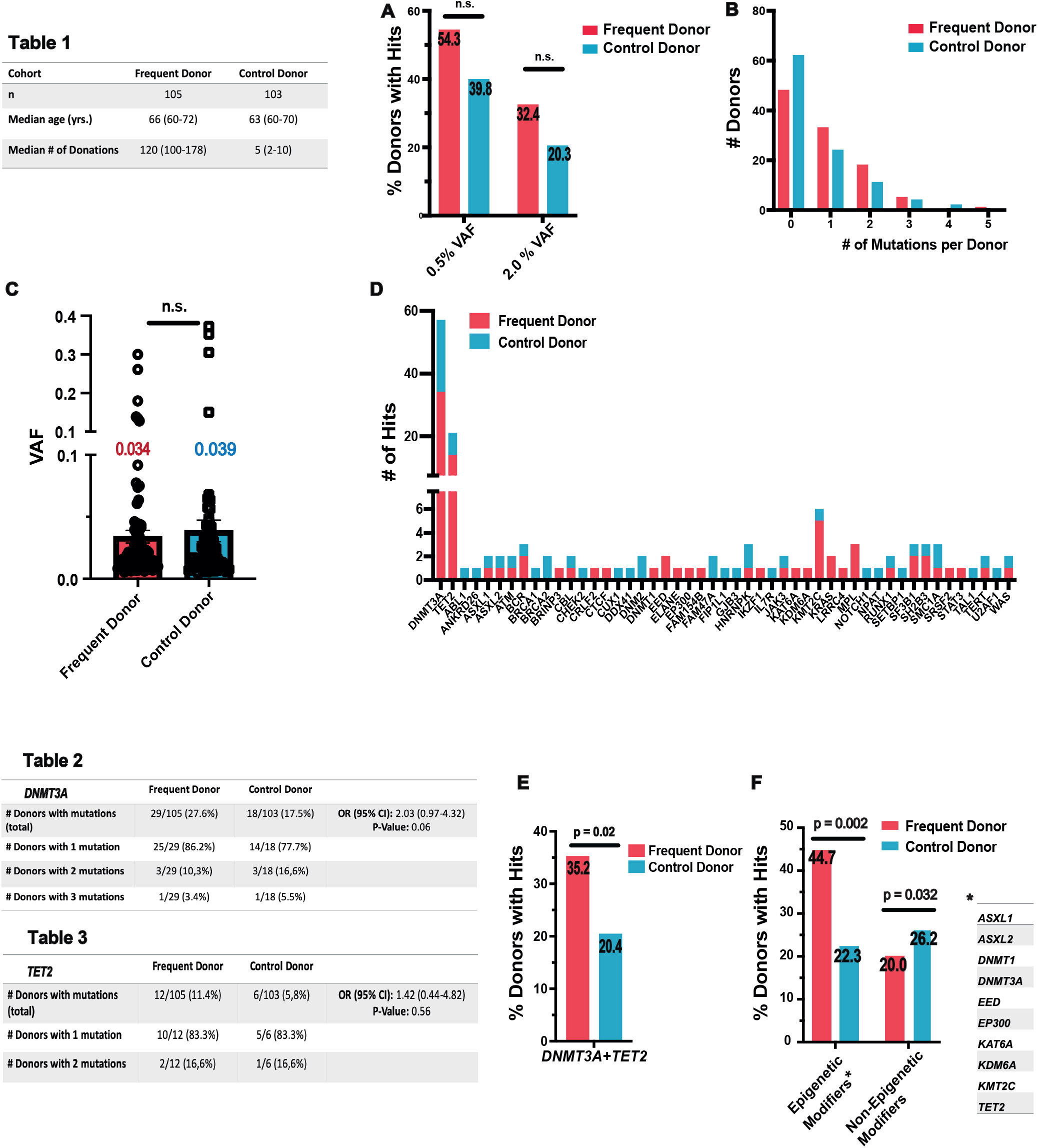
Mutations in epigenetic modifiers including *DNMT3A* and *TET2* are more prevalent in frequent blood donors. -Table 1 summarizes donor characteristics of the frequent donor (FD) and control donor (CD) cohort. (A) Percentage of donors with somatic mutations (Hits) in at least one of the 141 analyzed genes within the FD and CD cohort. The cutoff for the VAF (clone size) was set to 0.005 (0.5%). Analysis with a 0.02 (2%) cutoff is shown for comparison. Percentage values are indicated on the bars. For VAF cutoff 0.005: adjusted OR/CI:1.4 / 0.75-2.6, p=0.285. For VAF cutoff 0.02: adjusted OR/CI:1.25 / 0.61-2.55, p=0.538. (B) Number of donors with CH at VAF ≥0.005 with the numbers of mutations detected per donor. (C) VAF of all CH variants in the FD vs. CD detected at VAF ≥0.005. Mean values are specified. p=0.301. (D) All variants observed at a VAF ≥ 0.005, colored by cohort. Except for *DNMT3A* and *TET2* (most frequently detected variants), genes were ordered alphabetically. -Tables 2 and 3 show the prevalence and distribution of *DNMT3A* and *TET2* variants detected at a VAF ≥ 0.005. (E) Percentage of donors with a mutation in the *DNMT3A* and/or *TET2* gene detected at a VAF ≥ 0.005. Adjusted OR/CI:2.24 / 1.12-4.48, p=0.023. (F) Percentage of donors with somatic mutations (Hits) in one of the epigenetic modifier genes (* column on the right) and percentage of donors with somatic mutations in a non-epigenetic modifier. The cutoff for the VAF (clone size) was set to 0.005. Percentage values are indicated on the bars. Adjusted OR/CI:2.95 / 1.52-5.87, p=0.002 for epigenetic modifier genes; adjusted OR/CI:0.42 / 0.18-0.91, p=0.032 for non-epigenetic modifier genes.

CH variants with a VAF greater than 0.5% were detected in 57/105 (54.2%) of the FDs and 41/103 (39.8%) of the CDs (adjusted odds ratio (OR)/ confidence interval (CI):1.4 / 0.75-2.6, p=0.285, Figure 1A, supplemental Table 2). In both groups, most individuals had CH with a single mutation, comprising 33/57 (57.9%) in the FD and 24/41 (58.5%) in the CD cohort (Figure 1B). The VAFs of all detected variants did not significantly differ between the two cohorts (Figure 1C). Within the FD cohort, the presence of mutations could not be attributed to differences in donation frequency as the median time between two successive whole blood donations was 118 days (73-162) for donors with mutations and 111 days (83-192) for donors without detected CH variants (data not shown). When applying a VAF cutoff of 2%, as was used in the original CH studies^17,19,20^, CH variants were detected in 34/105 (32.4%) and 21/103 (20.3%) of the FD and CD cohorts, respectively (adjusted OR/CI:1.25 / 0.61-2.55, p=0.538, Figure 1A). Thus, independent of the VAF cutoff used there was no significant difference in the overall CH prevalence nor in the VAF distribution between the cohorts. All subsequent analyses were performed using mutations detected at a VAF cutoff of 0.5%.

### Mutations in epigenetic modifiers including *DNMT3A* and *TET2* are more prevalent in frequent blood donors

Consistent with previously reported studies on healthy, aged individuals^17,19,20,33,34^, *DNMT3A* (Supplemental Table 3) and *TET2* (Supplemental Table 4) variants were most common in both our cohorts (Figure 1D). Mutations in both genes were more prevalent in the FD compared to the CD cohort. There were 34 *DNMT3A* mutations in 29/105 FDs (main cohort) vs. 23 mutations in 18/103 CDs (Figure 1/Table2), and there were 14 *TET2* mutations in 12/105 FD donors vs. 7 mutations in 6/103 CD donors (Figure 1/Table 3). Overall, whereas the distribution of the VAFs was very similar between the two groups (Supplemental Figure 1A), a significantly higher fraction of FD donors carried a mutation in *DNMT3A* or *TET2* compared to the individuals in the CD cohort (adjusted OR/CI: 2.24 / 1.12-4.58, p=0.023, Figure 1E).

When the analysis was extended to genes encoding epigenetic modifiers as a whole (*ASXL1, ASXL2, CREBBP, DNMT1, DNMT3A, EED, EP300, KAT6a, KDM6a, KMT2c and TET2*) the trend towards a higher prevalence of mutations within the FD was even more pronounced. Variants in an epigenetic modifier gene were detected in 44.7% vs. 22.3 % of FDs vs. CDs, respectively (adjusted OR/CI: 2.95 / 1.52-5.87, p=0.002, Figure 1F). By contrast, CH in the remainder of genes (non-epigenetic modulators) was detected in 21/105 (20.0%) and 27/103 (26.2%) of the FD and CD groups, respectively (adjusted OR/CI: 0.42/ 0.18-0.91, p=0.032, Figure 1F). A mutation in *DNMT3A* was significantly more likely (conditional OR: 11/1, p=0.006) to co-occur with a second hit in *DNMT3A* or *TET2* (in both cohorts combined) rather than an additional mutation occurring in a non *DNMT3A* / *TET2* epigenetic modifier (termed OTHER, Supplemental Figure 1B and C).

In summary, we found the overall trend towards higher prevalence of CH within the FD cohort to be driven by mutations in genes encoding for epigenetic modifiers, with *DNMT3A* and *TET2* being the most common.

### *DNMT3A* mutations enriched in frequent blood donors likely result in reduced enzyme activity

*DNMT3A* and *TET2* mutations were distributed throughout the length of the whole gene (Figure 2A) consistent with previous reports for CH associated mutations in these drivers^33^. The frequency of the acute myeloid leukemia (AML) hotspot mutations at position 882 in the *DNMT3A* gene^33,35,36^ was low and did not differ between the cohorts: 3/36 mutations in the extended FD cohort vs. 2/23 mutations in the CD cohort (Supplemental Table 3).

**Figure 2.**
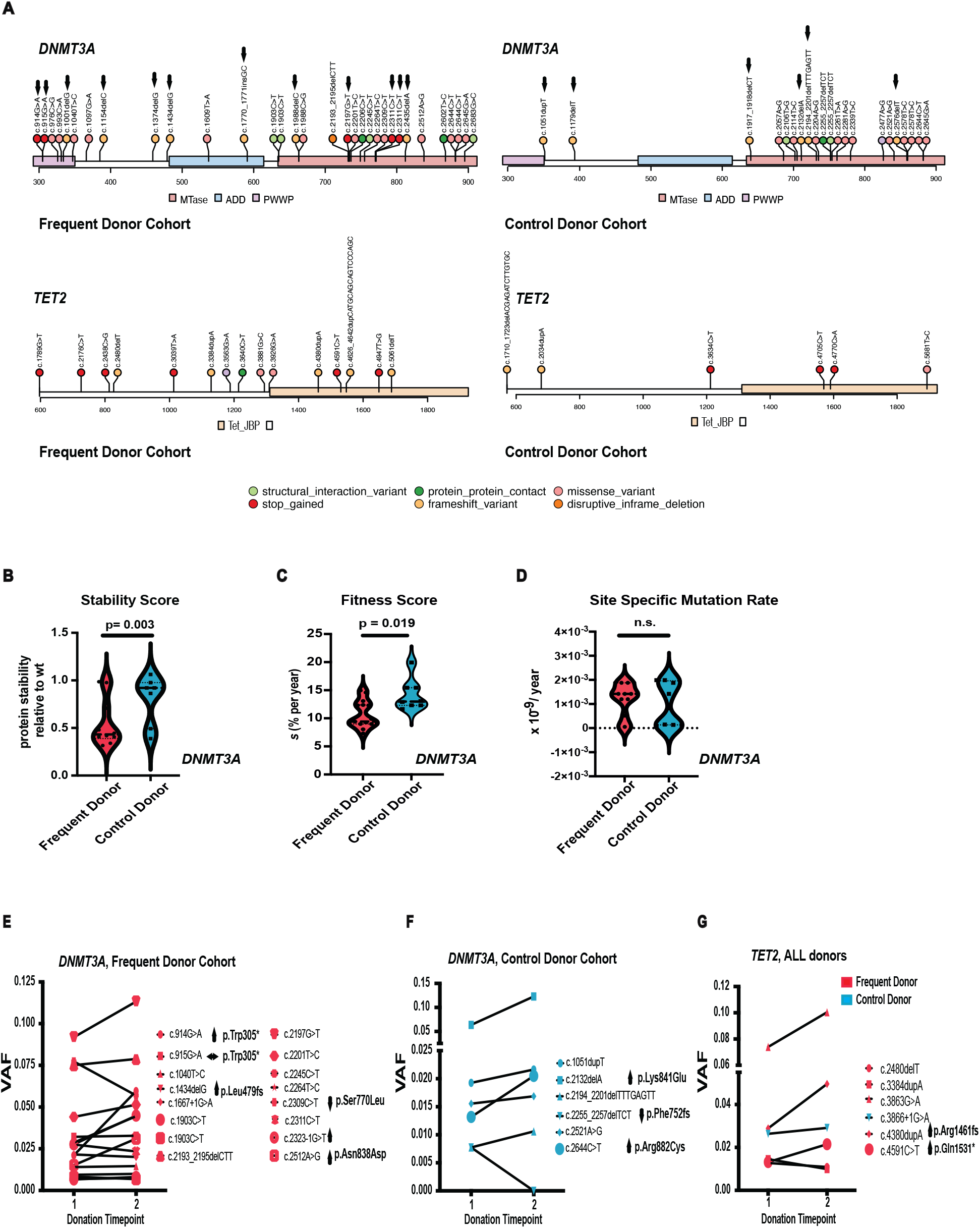
Mutations in frequent blood donors are not pathogenic. (A) Locations of the mutations in *DNMT3A* and *TET2* are shown (detected at a VAF ≥ 0.005, extended FD and main CD). The events are color-coded based on their effects on the protein (see bottom legend). For DNMT3A, the locations of the PWWP (proline–tryptophan– tryptophan–proline motif), the ADD (ATRX, DNMT3, and DNMT3L)-type zinc finger, and the methyltransferase (MTase) domains are shown. For TET2, the oxygenase domain of the 2OGFeDO superfamily (TET_JBP) is shown. All *TET2* mutations detected in either cohort as well as all *DNMT3A* mutations from the control cohort are depicted. Four splice site mutations in DNMT3A from the extended frequent donor cohort (FD) (c.855+2delT, c.1667+1G>A, c.2083-1G>A and c.2323-1G>T) are not shown. *DNMT3A* mutations resulting in premature translational stops are marked with arrows. See Supplemental Tables 2 and 3 for full lists of events. (B) Analysis of stability scores for *DNMT3A* mutations from the FD and CD cohort that were matched to the variants characterized by Huang et al.^42^ (See Supplemental Table 3), p=0.003. (C-D) Analysis of the fitness (C, p=0.019) and site-specific mutation rate (D) for *DNMT3A* mutations from the frequent and control donor cohort that were matched to the variants characterized by Watson et al.^33^ (See Supplemental Table 5). (E-G) Longitudinal analysis of variants. Samples were collected at a second timepoint (approximately 1 year later, see Supplemental Tables 6 and 7 for details). 16 *DNMT3A* variants from the FD (E) and 6 *DNMT3A* variants from the CD cohorts (F) were analyzed. A VAF of 0.000 for the second donation of Donor #444 indicates that the mutant allele was not detected at the set VAF cutoff of 0.005. However, a VAF of 0.002 was determined using the actual read count ratio in this sample. Variants that showed a change in the VAF between the two time points are specified in the graphs. The two W305* variants analyzed displayed very different kinetics: one VAF increased from 0.021 to 0.059 (Donor #8, Extended FD) whereas the other one remained unchanged (VAF of 0.032 at both time points, Donor #3). All six TET2 mutations analyzed are shown in panel G, colored by the group the donor belongs to.

Interestingly, we observed a trend towards a higher fraction of the *DNMT3A* variants from the FD cohort being or resulting in a premature translational stop (12 out of 36 within the extended FD cohort) compared to the control cohort (6 out of 23). Three of these FD DNMT3A nonsense variants, W305*, S663fs and E733*, were chosen for structural modelling in comparison to full length DNMT3A (Supplemental Figure 1D). The mutations were chosen based on the type of mutation as well as their VAF of > 5 % likely indicative of a selective advantage. The early stop variant W305* (Exon 8 out of 23) is predicted to be degraded due to nonsense-mediated mRNA decay (NMD)^37,38^. If translated, the corresponding truncated protein lacks 2/3 of its length including the entire methyltransferase domain. The S663 frameshift mutation results in a premature stop in exon 18 (amino acid position 704) and is therefore likely to be also degraded via the NMD pathway^39^. In case of NMD escape and effective translation, the DNMT3A S663fs protein lacks key residues required for DNA binding such as the target recognition domain (TRD) (residues R831–F848), the catalytic loop (residues G707–K721) as well as the DNMT3A–DNMT3A homodimer interface residues (S881, R882, L883, and R887)^37,38^. In addition to its reduced methyltransferase activity, DNMT3A S663fs is therefore predicted to be incapable of complexing with wild-type (wt) DNMT3A protein and sequestering the latter, as has been reported for the R882 variant^40^. Lastly, the E733* variant with a premature stop in exon 19 out of 23 is also expected to be subject to NMD. Upon translation, the E733* DNMT3A protein will contain the catalytic loop residues yet lack the TRD along with the homodimer binding capacity. Thus, compared to previously described mutations capable of interacting with wt DNMT3A protein, including the AML hotspot variant R882H, the three FD variants W305*, S663fs (704*), F733* are likely to cause a “simple” reduction in DNA methylation levels rather than aberrant methylation patterns associated with R882H^37,41^. In line with this, when using the recently introduced stability score^42^ to characterize the DNMT3A variants from both cohorts, the scores of the variants matched from the FD cohort were significantly lower compared to the scores of the CD cohort variants (p=0.003, Supplemental Table 3 and Figure 2B). Lower stability described by Huang et al. was directly linked to regulated degradation of the DNMT3A protein, which again is reflected in reduced rather than aberrant enzyme activity^42^. Taken together, both, *in silico* structural predictions as well as previously reported functional characterization suggest an enrichment of DNMT3A variants with reduced enzyme activity in the FD compared to the CD.

### Mutations in frequent blood donors are not pathogenic

We next compared the impact of the detected variants using a tool that scores variants with regard to their functional impact, deleteriousness and pathogenicity. The Combined Annotation Dependent Depletion (CADD)^43-45^ tool has previously been used to score CH variants^46^ and was applied to mutations found in our cohorts. We did not find significant differences between the CADD scores of FD vs. CD cohorts when considering all mutations or individual *DNMT3A* or *TET2* mutations (Supplemental Figure 1E-G).

When using the fitness score (*s*) introduced by *Watson et al*., which is based on a retrospective analysis of the large-scale CH studies published over the past decade^33^, we were able to match 16 *DNMT3A* mutations found in the FD and 8 *DNMT3A* mutations from the CD to those reported and analyzed in that study (see Supplemental table 5 for the list of “overlapping” mutations). *S* values of 4-10% per year have been categorized as moderate, whereas s values higher than 10% per year indicate that a mutation is likely to take over the bone marrow (BM) and displace non-mutated hematopoiesis^33^. Interestingly, the mean *s* was 10.6% per year for the FD compared to 14.1% per year for the CD *DNMT3A* variants (p=0.019, Figure 2C). The high frequency of the FD variants despite a lower fitness suggests that the mutant clones experienced a specific environmental growth stimulus under frequent blood donations to reach their observed sizes and frequencies. There was no difference in the site mutation rate values, a second critical parameter contributing to the penetrance of a variant (Figure 2D).

Thus, the higher frequency of certain hits observed in FDs was not associated with a higher fraction of mutations predicted to be pathogenic according to the CADD scoring system. In fact, the fitness advantage score-based comparison of the two cohorts even suggests an enrichment for variants with a lower fitness in the frequent donor as compared to the control donor cohort.

### *DNMT3A* mutations are acquired early in life

For selected donors and *DNMT3A* variants, we performed a more comprehensive analysis using digital droplet PCR (ddPCR). At the second timepoint of sample collection, in addition to whole PBMCs, several mature cell fractions (B-, T-cells and monocytes) were analyzed along with the immature CD34+ HSPC compartment. The early stop variant W305* was chosen as it was present in two donors (donor #3 and #8). Two additional mutations, E733* and F732del from donors #42 and #31, respectively, were chosen because of their high VAFs. The second, smaller clone found in donor #42 (L347P) was also analyzed as well as the well-known preleukemic R882C clone detected in donor #371. The changes in VAF as detected using error-corrected deep sequencing were verified using ddPCR analysis of whole PBMC samples (Supplemental Figure 2A). In line with previous studies^47-50^, all five mutations analyzed were detected in all four of the sorted populations, including in T cells, pointing towards their acquisition and selection at the level of the multipotent HSC, and likely decades ago (Supplemental Figure 2B-F). This observation also fit with the demonstration that *DNMT3A* mutations occur and expand rather early in life^51^.

### Majority of the clones remain stable with time

Analyses of the kinetic behavior of variants’ expansion using longitudinal sampling has been shown to be particularly informative when trying to predict their malignant potential^51-55^. Having identified donors of interest carrying CH, we were able to obtain at least one additional consecutive donation sample from 16 donors from the extended FD and from 6 of the CD cohort. The difference in sample number is explained by the different donation habits. PB leukocytes were collected when the donors’ next blood donation sample became available, as they were pseudonymized to the investigators and could not be individually contacted for additional blood samples. Median time between the two timepoints analyzed was 364 (218-512) and 411 (245-520) days, for the FD and CD cohorts, respectively. 27 out of the 28 mutations analyzed longitudinally were in *DNMT3A* or *TET2*. All mutations were detected at both timepoints. The VAF of the majority of *DNMT3A* mutations did not change between the two time points analyzed (Figures 2E-F). This is consistent with a recent report demonstrating rapid expansion of *DNMT3A*-mutant clones early, but rather slow growth later in life^51^. Nevertheless, within the frequent donor cohort, 4 out of 16 *DNMT3A* mutations analyzed displayed a 1.5-3-fold increase in their VAF (Figure 2E and Supplemental Table 6). Interestingly, the same early stop W305* mutation detected in two different donors (both in FD) displayed different growth patterns. In donor #3 (W305*/ c.915G>A) the VAF remained stable over a period of one year (399 days). This was true for all three *DNMT3A* variants detected in this donor. By contrast, in donor #8 (W305*/ c.914 G>A, extended FD cohort, see Methods for detailed description) the *DNMT3A* clone expanded almost 3-fold. Within the CD cohort, 2 out of 6 DNMT3A mutations displayed an approximately two-fold increased VAF at the later timepoint (Supplemental Table 6); one of them was the well-characterized preleukemic mutation R882C (Figure 2F).

Of the five *TET2* mutations from the FD, two displayed an increase in their VAF at the later timepoint, one decreased slightly and two remained stable (Figure 2G). The only *TET2* mutation from the CD showed a similar VAF at the second timepoint (Figure 2G). In line with this, *TET2* mutated clones have previously been demonstrated to grow uniformly across ages^51^.

### *DNMT3A* mutations from frequent donors expand in response to erythropoietin but not to inflammatory stimuli

One of the first and most immediate responses of the body following whole blood donation and its associated drop in red blood cells (RBC) and hemoglobin by 12 and 10 %, respectively^56^, is secretion of erythropoietin (EPO) by the kidneys to stimulate erythropoiesis in the BM for rapid replenishment of the lost red blood cells^57^. Accordingly, up to two-fold increased concentrations of EPO are detected in the serum of blood donors for up to 56 days following a single donation^57^. We sought to functionally link the presence of variants detected in frequent donors with their responsiveness to stimulation with EPO.

As described above, nonsense *DNMT3A* mutations were enriched in the FD cohort. The three *DNMT3A* mutations subjected to structural analysis based on the type of mutation as well as their VAF were also chosen for *in vitro* analysis: W305*, S663fs (704*) and E733*. Human HSPCs, isolated from umbilical cord Blood (CB) or adult BM, were edited using CRISPR/Cas9 mediated homology directed repair (HDR) to introduce the chosen mutations. Following editing, all HSPCs cells were cultured using long-term culture-initiating cell (LTC-IC) assays in the presence or absence of EPO or inflammatory challenges. The VAF of the introduced variants was assessed after five weeks of culture using next generation sequencing (Figure 3A). Two different concentrations of EPO were tested; both promoted robust erythroid differentiation of HSPCs as observed by the expansion of CD235a+CD71+ cells (Figure 3B-C). All three mutations supported 2-3-fold expansion in response to EPO but not in response to LPS treatment (Figure 3D). Moreover, the erythroid fraction of the culture (if present) showed a higher VAF of the introduced mutations compared to the non-erythroid cells (Figure 3E), indicating their selection under erythropoietic stimulation.

**Figure 3.**
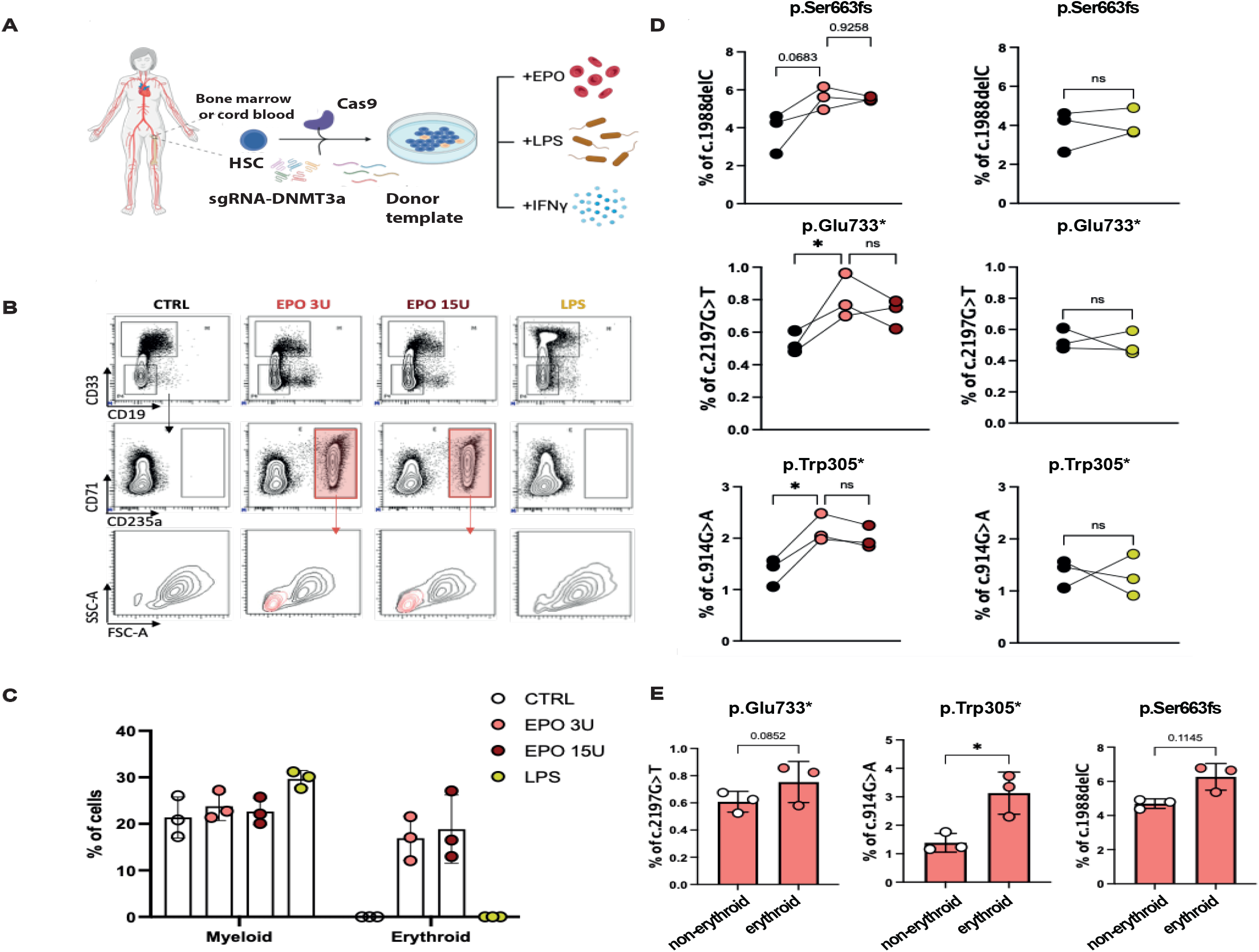
Erythropoiesis-induced stress expands DNMT3A-mutant clones found in frequent blood donors. (A) Schematic representation of genetic engineering of human HSCs to introduce mutations found in frequent blood donors and perform LTC-IC in the presence of different stimuli. (B) Representative flow cytometric analysis after 5 weeks in the presence of EPO or LPS and (C) quantification of the non-erythroid (CD33^+^) and erythroid (CD235a^+^CD71^dim/high^) myeloid lineages found in the different conditions. (D) Sequencing results for each mutant clone, showing expansion upon EPO treatment (left) but no differences upon LPS treatment (right). (E) Comparison of the presence of each mutant clone between non-erythroid (CD235a^-^CD71^-^) and erythroid (CD235a^+^CD71^dim/high^) populations in the EPO 3U/ml condition. Each dot represents an independent biological donor. Paired t-test for each biological donor was used for the statistical significance of the percentage of the DNMT3A-mutant clones between different conditions. *, p < 0.05.

In an additional set of experiments, R882C and R882H mutated CB HSPCs were engineered and the behavior of these preleukemic variants was compared with the three non-R882 variants from the FDs. Preleukemic *DNMT3A* clones had been reported to expand upon chronic IFNγ treatment^21^. Indeed, our data confirmed this with both R882 clones being highly responsive to IFNγ but without expansion when treated with EPO (Figure 4A-B). Interestingly, while the modified cells carrying the FD variants again expanded upon EPO treatment, they did not show expansion in response to chronic inflammatory challenges like LPS or IFNγ (Figure 4A-B). Similar trends were observed with edited CD34+ cells isolated from the BM, although the variability was much higher (Supplemental Figure 3). These results indicate that different *DNMT3A* variants can introduce a distinctive selective advantage to blood cells and therefore be indicative of the environmental signals the cells have been exposed to. Clones with preleukemic mutations are selected in inflammatory environments. Meanwhile the *DNMT3A* clones enriched in frequent blood donors are selected in erythropoietin-rich environments and fail to respond to inflammatory cues.

**Figure 4.**
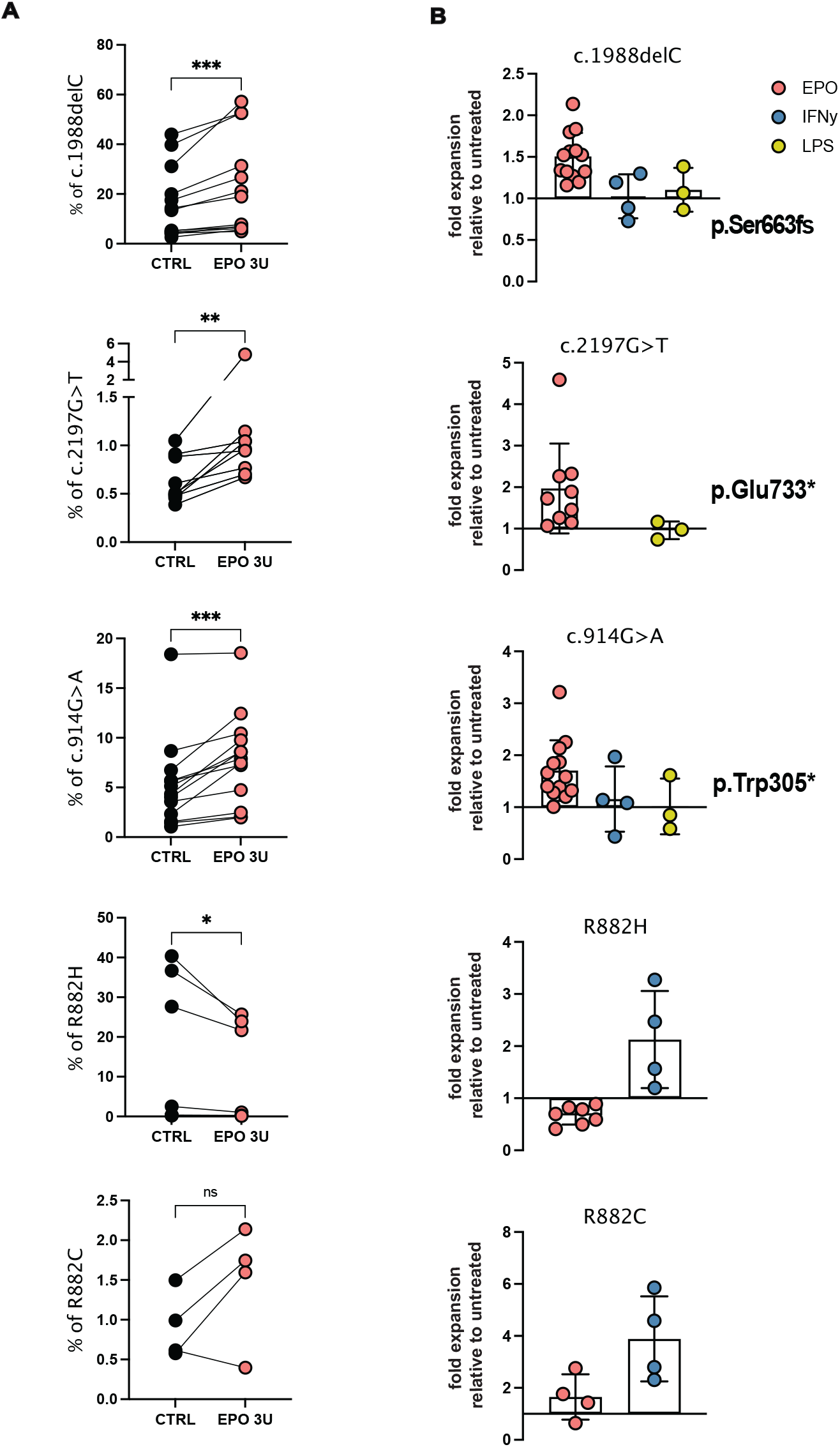
Non-malignant *DNMT3A*-clones associated with blood donation expand in EPO-induced stress while preleukaemic R882-mutant clones expand in IFNγ-induced stress. Sequencing results of all cord blood donors tested (n = 4-13) for each mutant clone in percentage (A) or fold-change expansion to untreated condition (B). For clone p.Ser663fs and clone p.Trp305* 13 biological donors were tested over 4 independent experiments. For clone p.Glu733* 9 biological donors were tested over 3 independent experiments. For clone R882C 4 biological donors were tested in 1 experiment. For clone R882H 7 biological donors were tested in 2 independent experiments. Each dot represents an independent biological donor. Paired t-test for each biological donor between different conditions was used for the statistical significance of the percentage of the DNMT3A-mutant clones. *, p < 0.05; **, p < 0.01; *** p < 0.001

## Discussion

Considering how essential a steady blood supply is for modern surgery and medicine, the scarcity of studies exploring the long-term safety of blood donation is surprising. A number of studies have pointed towards an association between clonal hematopoiesis and red blood cell disorders such as anemia^29,30,32^ and erythrocytosis^31^. However, the potential effects of donation-associated erythropoietic stress on hematopoiesis have not been studied. To our knowledge, this is the first work looking at the effect of frequent blood donations on healthy clonal dynamics of the hematopoietic system.

Older male blood donors were chosen for the study due to a higher limit of maximum donations per year for male compared to female donors^2^. We found no significant increase in prevalence of CH in frequent donors (median of 120 donations) compared to age-matched control donors (median of 5 donations). With a life span of 120 days, all RBCs are replaced three times per year^58^. Each donated unit adds 2.5-5% excess erythropoietic stress in that year, a situation which likely favors the outgrowth of variants responding to erythropoietin stimulation. This may explain the higher prevalence of epigenetic modifier-driven CH detected in frequent blood donors. The VAFs of detected variants were not significantly different between the two groups and are therefore likely primarily determined by the age of the donor or other, still unknown factors. The CH prevalence of 20.3% detected in the control donors is higher than the numbers reported in previous studies for similar age groups in which CH was reported in 5-10% of individuals at a VAF cutoff of 2%^17,19,20^. This is likely due to the differences in the sensitivity of sequencing panels used and the fact that, in contrast to previously studied mixed cohorts, our control cohort consisted exclusively of male individuals.

In agreement with most aging-associated CH studies^17-20,33,34^, *DNMT3A* and *TET2* were the two most commonly mutated genes in both cohorts. The observed significant increase in donors carrying a hit in *DNMT3A* and/or *TET2* was even more pronounced when genes encoding for epigenetic modifiers were considered as a class. These are typically considered to be the so-called “early hits” and deemed “neutral” with regard to their malignant potential^48,59-62^. Hence, we did not find any differences in the pathogenicity scores of the variants detected in both groups. This important observation is consistent with a recent report that found, if anything, a reduced risk of hematologic malignancies associated with blood donations^8^. Interestingly, when the recently described fitness score (*s*)^*33*^ was applied, the *DNMT3A* variants identified in the frequent donor cohort had a significantly lower score compared to the ones found in the control group. This suggests that mutations selected upon bleeding associated stress, which we assume to be overrepresented in our test cohort, display moderate fitness in the general population and in the absence of additional stimuli. Accordingly, our analysis of consecutive samples confirmed that the VAF of the majority of *DNMT3A* mutations remains stable. This is consistent with a recent long-term follow-up study showing that *DNMT3A* mutations are more likely to expand early in life and display a slower growth in older age^51^.

The current paradigm in the field is that specific cues from the environment are necessary to favor the outgrowth of HSPCs carrying mutations they acquire during the course of life^24,25,51,63^. Different environmental stress signals will have different impact on the fate of the diverse mutant clones^21,64-68^. Understanding how specific stress factors promote the expansion of CH clones carrying certain mutations may help us prevent the development of hematological malignancies and the pathologies associated with CH. Blood donation associated cell loss represents a stress signal that initiates changes in the proliferative behavior of HSPCs and their differentiation toward erythrocytes^69,70^. Consistent with this, we recreated this stress signal *in vitro* by treating HSPCs with EPO. We also analyzed whether the *DNMT3A* clones found in frequent blood donors are selected upon inflammatory stress signals which are known to favor the outgrowth of preleukemic *DNMT3A* clones (R882H and R882C)^15,21^. The observed selectiveness of HSPC responses to different stress stimuli with *DNMT3A* mutations from the frequent blood donors, which expanded upon exposure to EPO but not upon inflammatory signals, and the inverse observation made for R882 mutated cells, indicates that there is indeed an enrichment of EPO responsive clones that occurs in frequent donors and these clones behave differently from the known preleukemic *DNMT3A* clones. Functional differences between different *DNMT3A* mutations are well known and have been comprehensively studied^42,71,72^. Thus, a new class of *DNMT3A* mutational effects, namely those resulting in decreased protein stability, has been defined and functionally validated and is of particular interest in comparison to the dominant-negative, stable DNMT3A variants such as R882H^42^. Reduced DNMT3A protein levels of EPO responsive variants were predicted using *in silico* analysis. Moreover, the average stability score^42^ of FD DNMT3A variants was significantly lower compared to that of CD DNMT3A variants. A phenotype of reduced DNMT3A activity enriched among FD variants as opposed to aberrant methylation associated in particular with the preleukemic R882 variants would explain their different responsiveness upon exposure to different cytokines.

Analysis of lineage contribution for selected *DNMT3A* variants revealed similarities with the previously described phenotype of *DNMT3A* mutated hematopoiesis^47,66,73,74^. We detected higher VAF of all but one of the analyzed *DNMT3A* mutations in the myeloid as compared to lymphoid lineage cells isolated from the donors. Accordingly, an increase in granulocytic lineage at the expense of the lymphocytic one has been well described in individuals suffering from Tatton-Brown-Rahman syndrome, an autosomal-dominant disorder caused by germline mutations in *DNMT3A* as well as in *DNMT3A* knockout mice (KO)^50,74^. More recently, single cell transcriptome and methylome analysis of DNMT3A-KO mice point towards the expansion and transcriptional bias to an erythro-megakaryocytic fate^47,66^. Overall, it appears that the different types of *DNMT3A* mutations share a proneness for myelo-erythropoietic skewing of HSPCs. Indeed, HSCs do express the EPO-receptor and differentiation trajectory directly from HSCs towards megakaryocyte-erythroid progenitors has been described^75-78^.

Our comprehensive analysis of aged blood donors shows that frequent blood donors have a higher prevalence of mutations in genes encoding for epigenetic regulators, including the two most commonly mutated CH genes, *DNMT3A* and *TET2*. To our knowledge, this is the first CH study focusing on a cohort of overwhelmingly healthy individuals as opposed to looking for mutations associated with certain disease conditions. Importantly, none of the mutations previously linked with increased likelihood of developing hematologic malignancies were found enriched in the frequent donors, strongly suggesting that frequent blood donation not only does not predispose for hematological cancers but may potentially even be protective. The increased frequency of EPO-responsive variants in frequent donors suggests causality. While acquisition of a given variant is stochastic, microenvironment driven evolution under extensive blood donation, a previously undescribed type of selection pressure, appears to favor clones with low to moderate growth potential i.e. fitness. This phenomenon is an example of ongoing Darwinian evolution within a human stem cell pool in response to chronic stress, in this case mediated by frequent blood loss and exposure to elevated EPO levels. Importantly, responsiveness of HSPCs to EPO seems not to be associated with higher intrinsic fitness of the cells. Overall, our study raises no concerns about the safety of blood donation, since even decades of regular donations do not result in apparent pathologic changes of the clonal dynamics of HSPCs. Nevertheless, marks of erythropoietic stress are detected in the blood cell genomes of frequent blood donors in the form of higher frequency of epigenetic modifier driven CH and specific *DNMT3A* mutations rendering cells more EPO responsive, which we believe could be prevented if more eligible individuals took on the responsibility of blood donation.

## Methods

### Blood Donors

Blood donors were healthy male volunteers who donated at the German Red Cross Blood Service Baden-Württemberg-Hessen between December 2019 and June 2020 as well as December 2020 and November 2021 (for consecutive samples). All donors signed informed consent allowing for anonymous processing of samples. All donor IDs are research IDs introduced in course of the study for the purpose of data analysis exclusively and do not allow an identification of the donors to anyone outside the research group.

### Human HSCs

Umbilical Cord Blood (UCB) was obtained from full-term donors after informed consent at the Royal London Hospital (London, U.K.). Mononuclear cells (MNCs) were isolated by density centrifugation using Ficoll-Paque (GE 67 Healthcare). Anonymized human BM samples from consenting healthy volunteers were provided as approved by Ethics Committee vote #329/10. MNCs were depleted for lineage positive cells using an EasySep Human Progenitor Cell Enrichment Kit (Stem Cell Technologies) and HSPCs isolated as described previously^79^.

### Culture of *DNMT3A* mutant human HSCs under cellular stress

Human HSPCs (CD34^+^CD38^-^) were cultured in StemSpanSFEM (Stem Cell Technologies) with 100 ng/mL rhFLT-3L, 100 ng/mL rhSCF, and 100 ng/mL rhTPO for 48 hours. CRISPR editing was then performed with the NEON Transfection system (Thermo Fisher) using the indicated small guide RNAs and donor templates (Supplemental Table 9). For LTC assays, a MS-5 feeder-layer was seeded at 2×10^4^ cells/cm^2^ in a 12-well dish and irradiated (7.5 Gy) after 24 h. 6-12 h later, media was replaced with 1 ml of Myelocult H5100 (Stem Cell Technologies). 48 h following CRISPR-induced modification, 1,000 HSPCs were transferred to MS-5 plates. Once a week, 500 µL of media was replaced with the following stimuli: Erythropoietin (Peprotech; 3 or 15 U/mL as indicated), human IFN-γ (Bio-Techne; 100 ng/mL) or LPS (Sigma; 1 µg/mL). After 5 weeks, cells were collected and analyzed by flow cytometry. Different cell populations were sorted for DNA extraction.

Additional information on the methods used is provided in the **Supplementary Appendix**.

## Data Availability

All data produced in the present study are available upon reasonable request to the authors

## Acknowledgements

First and foremost, we thank all volunteer blood donors for their dedication and altruism with which they continue to donate life-saving resources!

The authors thank Stephanie Müller, the physicians and the technical staff involved in collection and processing of the whole blood units as well as the IT Department at the German Red Cross Blood Donation Service Baden-Württemberg-Hessen for their assistance in identifying and supplying the samples. The authors thank Steffen Schmitt, Marcus Eich, Klaus Hexel, Tobias Rubner and Florian Blum from the German Cancer Research Center (DKFZ) Flow Cytometry Core Facility as well as the DKFZ Genomics and Proteomics Core Facility for their assistance. This work was partly supported by the SPP2036 and SFB873 funded by the Deutsche Forschungsgemeinschaft; the DKTK joint funding project “RiskY-AML”; the “Integrate-TN” Consortium funded by the Deutsche Krebshilfe and the Dietmar Hopp Foundation (A.T.). This work was partly supported by Cancer Research UK (FC001115), the UK Medical Research Council (FC001115), the Wellcome Trust (FC001115) to DB. H-HE was supported by the Kay Kendall fellowship (KKL1397). For the purpose of Open Access, the authors have applied a CC BY public copyright license to any Author Accepted Manuscript version arising from this submission.

We kindly thank Moritz Gerstung (German Cancer Research Center [DKFZ] and DKFZ-ZMBH Alliance) for critical revision of the manuscript, helpful comments, and discussion.

The authors declare no conflict of interest.

## Contributions

K.D., HE.H., D.E., K.I., L.A-M., P.A. performed experiments. K.D., H E.H., L.A-M., P.J. analyzed data; K.I. performed structural modelling; C. S. and K-S. A. performed statistical analysis. S.E. provided critical reagents for the study. K.D., HE.H., B.H., and T.A. designed the research; K.D., W.TN., B.D., B.H. and T.A. supervised the study. K.D., HE.H., W.TN., B.H., and T.A. wrote the original draft; K.D., HE.H., D.E., L.A-M., G.M., W.TN., B.D., B.H. and T.A. reviewed and edited the manuscript. All authors discussed, commented on, and approved the final version of the manuscript.

