## SupplementalAppendix for "Frequent whole blood donations select for DNMT3A variants mediating enhanced response to erythropoietin"

| | | |
|---|---|---|
| Supplemental Figure 1 | **Clonal hematopoiesis found in frequent blood donors is enriched in genes encoding for epigenetic regulators.** | **p.2** |
| Supplemental Figure 2 | *DNMT3A* variants detected in blood donors are present in the myeloid and lymphoid lineage. | p.3 |
| Supplemental Figure 3 | Bone marrow HSPCs harboring R882 mutations expand in IFNγ-induced stress while non-preleukaemic *DNMT3A*-clones expand in EPO-induced stress. | p.4 |
| Supplemental Figure Legends | | p.5 |
| Supplemental Table 1 | Blood Donor Cohorts Metadata | pp.6-8 |
| Supplemental Table 2. | List of all detected mutations | pp.9-11 |
| Supplemental Table 3. | List of all *DNMT3A* mutations | p.12 |
| Supplemental Table 4. | List of all *TET2* mutations | p.13 |
| Supplemental Table 5. | DNMT3A fitness scores and site-specific mutation rates | p.14 |
| Supplemental Table 6. | Longitudinal analysis | p.15 |
| Supplemental Table 7. | Differential blood count analysis of Buffy Coats | p.16 |
| Supplemental Table 8. | Digital droplet PCR Assays | p.17 |
| Supplemental Table 9. | CRISPR guides and HDR donor templates | p.18 |
| Supplemental Methods | | pp.19-23 |
| Supplemental References | | p.24 |

### Supplemental Figure 1

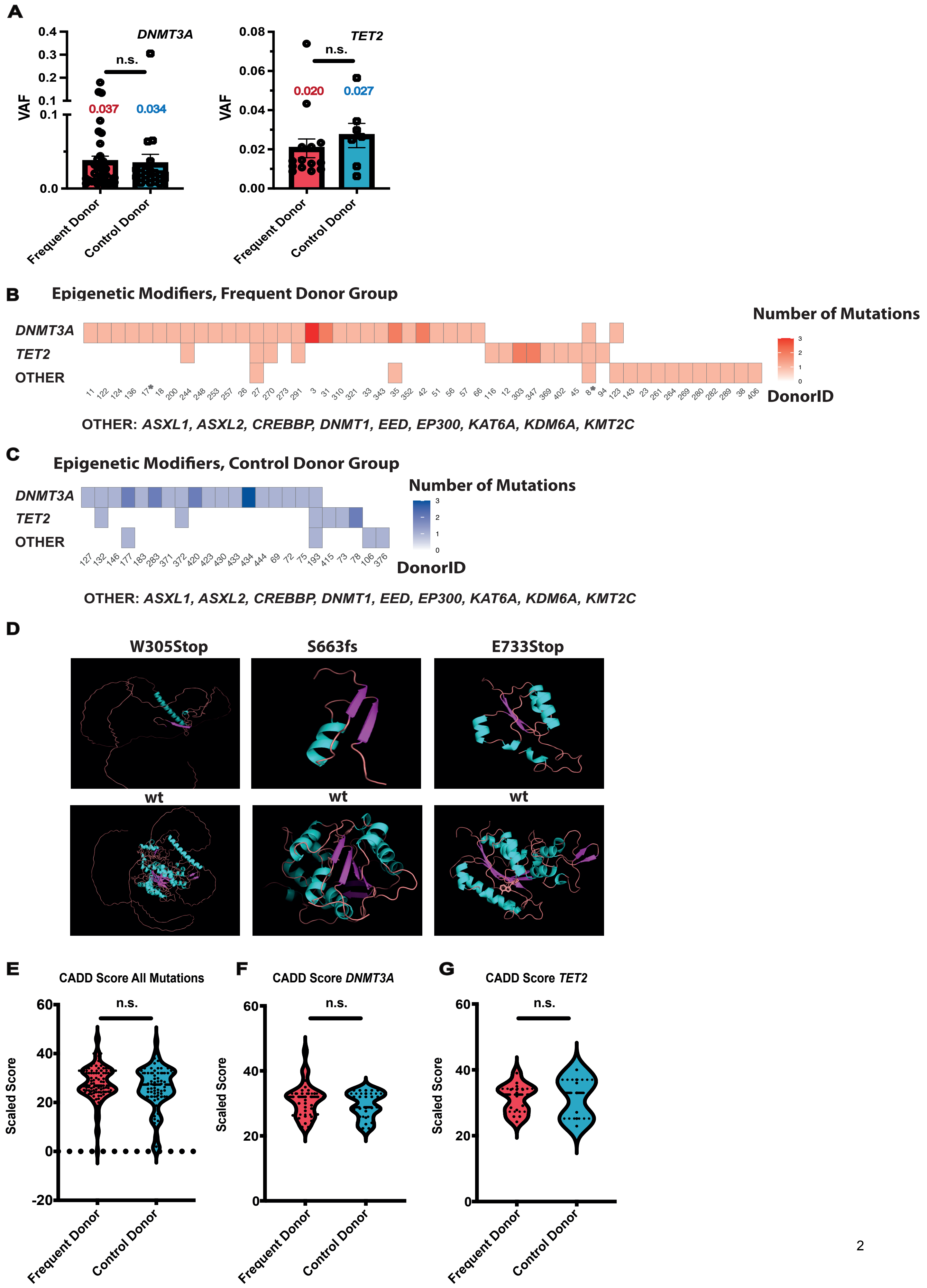

### Supplemental Figure 2

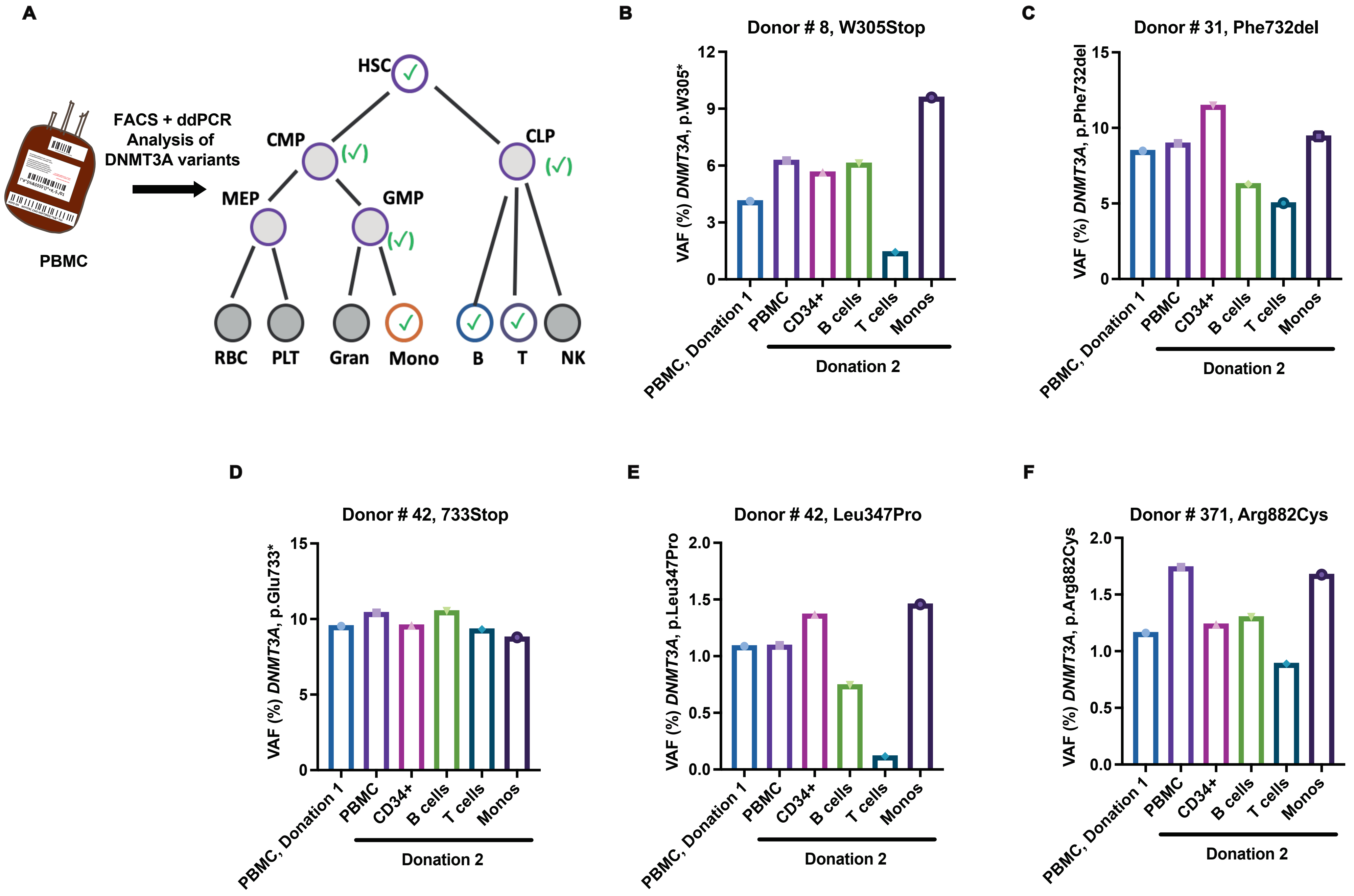

### Supplemental Figure 3

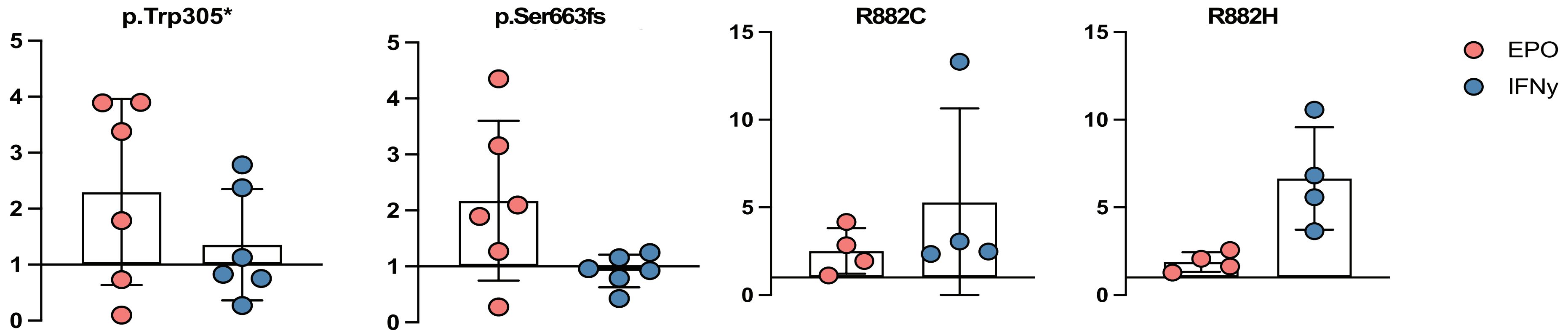

#### Supplemental Figure Legends

**Supplemental Figure 1. Clonal hematopoiesis found in frequent blood donors is enriched in genes encoding for epigenetic regulators.**

(A) VAF of all *DNMT3A* (left) and *TET2* (right) variants in the frequent (FD) vs. control donor cohort (CD) detected at a VAF ≥ 0.005. Mean values are specified above the bars. (B-C) Co-mutational analysis of the variants in epigenetic regulators detected in the (extended) FD (B) and CD (C) at a VAF ≥ 0.005. For each donor number and identity (*DNMT3A* vs. *TET2* vs. one of the following genes: *ASXL1, ASXL2, CREBBP, DNMT1, EED, EP300, KAT6A, KDM6A* or *KMT2C* (termed OTHER)) are depicted. Upon presence of one DNMT3A mutation a conditional odds ratio of 11/1 for having a second hit in *DNMT3A* or *TET2* as compared to OTHER was determined (p=0.006 based on McNemar exact test).

(D) Structural models of the DNMT3A variants W305*, S663fs (704*) and E733* in direct comparison to the wt protein were generated using homology modelling on SWISS-MODEL[83,84] based on the crystal structure of DNMT3A available on PDB under the alias 5YX2[37].

(E-G) Combined Annotation Dependent Depletion (CADD) based scoring[44,45] of the CH variants detected at a VAF ≥ 0.005 was performed. Scaled score for all (p=0.476), *DNMT3A* (p=0.358) and *TET2* (p=0.725) variants are shown in panel E, F and G, respectively.

**Supplemental Figure 2. *DNMT3A* variants detected in blood donors are present in the myeloid and lymphoid lineage.**

(A) Schematic presentation of analysis of mature and immature cell fractions isolated using FACS (green check mark) from PBMC of the frequent blood donors Donor 8 (B), 31, (C), 42 (D and E) and the control blood donor 371. VAF of each mutation was analyzed using digital droplet PCR (ddPCR) performed concurrently on the whole PBMC samples (timepoint 1 and 2) as well as the CD34+ cells, T- and B-cells and monocytes collected at the second donation timepoint. All five variants tested were detected in all 4 sorted cell populations, indicative of their presence in the myeloid (CMP, GMP, green mark in brackets) and lymphoid (CLP, green mark in brackets) lineage.

**Supplemental Figure 3. Bone marrow HSPCs harboring R882 mutations expand in IFNγ-induced stress while non-preleukaemic *DNMT3A*-clones expand in EPO-induced stress.**

Fold-change expansion (treated, with EPO or IFN***γ***, compared to untreated condition) of different mutations introduced in bone marrow HSPCs (4-6 biological donors tested). Each dot represents an independent biological donor. For each biological donor, a paired t-test was used to compare the percentage of the *DNMT3A*-mutant clones between different conditions

#### Supplemental Table 1. Blood Donor Cohorts Metadata

| Group | DonorID | NumberOfDonations | SequencingDepth |
|---|---|---|---|
| FD | 2 | 144 | 965.39 |
| FD | 3 | 115 | 1248.82 |
| FD | 4 | 120 | 958.07 |
| FD | 7 | 105 | 684.22 |
| FD | 9 | 133 | 664.41 |
| FD | 10 | 125 | 844.18 |
| FD | 11 | 112 | 805.95 |
| FD | 12 | 176 | 1016.42 |
| FD | 18 | 104 | 963.99 |
| FD | 19 | 101 | 2114.96 |
| FD | 20 | 136 | 853.85 |
| FD | 21 | 101 | 834.39 |
| FD | 22 | 110 | 1071.38 |
| FD | 23 | 126 | 633.68 |
| FD | 26 | 153 | 639.95 |
| FD | 27 | 107 | 809.51 |
| FD | 29 | 109 | 842.99 |
| FD | 30 | 110 | 715.83 |
| FD | 31 | 138 | 699.5 |
| FD | 33 | 105 | 431.14 |
| FD | 34 | 126 | 698.1 |
| FD | 35 | 104 | 873.91 |
| FD | 36 | 101 | 769.25 |
| FD | 38 | 151 | 1451.24 |
| FD | 40 | 154 | 1155.28 |
| FD | 41 | 167 | 626.71 |
| FD | 42 | 124 | 774.9 |
| FD | 44 | 146 | 855.66 |
| FD | 45 | 101 | 802.71 |
| FD | 46 | 125 | 804 |
| FD | 48 | 100 | 884.29 |
| FD | 50 | 127 | 536.82 |
| FD | 51 | 122 | 751.87 |
| FD | 53 | 101 | 692.95 |
| FD | 56 | 119 | 720.45 |
| FD | 57 | 122 | 1056.89 |
| FD | 66 | 102 | 1045.96 |
| FD | 94 | 161 | 1019.4 |
| FD | 116 | 102 | 1181.78 |
| FD | 122 | 107 | 1837.21 |
| FD | 123 | 115 | 1400.59 |
| FD | 124 | 128 | 1628.78 |
| FD | 126 | 130 | 1169.06 |
| FD | 136 | 111 | 1308.2 |
| FD | 143 | 134 | 1481.45 |
| FD | 147 | 140 | 901.08 |
| FD | 150 | 110 | 2003.89 |
| FD | 152 | 131 | 1473.09 |
| FD | 154 | 102 | 973 |
| FD | 167 | 101 | 1077 |
| FD | 190 | 163 | 1064.58 |
| FD | 200 | 111 | 1432.01 |
| FD | 208 | 137 | 1369.68 |
| FD | 224 | 126 | 1440.35 |
| FD | 231 | 101 | 1470.76 |
| FD | 239 | 126 | 1385.89 |
| FD | 244 | 121 | 2061.18 |
| FD | 245 | 106 | 1501.18 |
| FD | 248 | 111 | 919.07 |
| FD | 253 | 103 | 1277.43 |
| FD | 256 | 103 | 1586.7 |
| FD | 257 | 125 | 1202.03 |
| FD | 261 | 149 | 1070.68 |
| FD | 264 | 172 | 1048.1 |
| FD | 266 | 140 | 1024.56 |
| FD | 268 | 132 | 1024.04 |
| FD | 269 | 109 | 1211.45 |
| FD | 270 | 100 | 1088.81 |
| FD | 271 | 120 | 952.81 |
| FD | 273 | 135 | 1548.4 |
| FD | 274 | 128 | 1061.47 |
| FD | 278 | 111 | 2289.06 |

**Supplemental Table 1. Blood Donor Cohorts Metadata**

| | | | |
|---|---|---|---|
| FD | 280 | 133 | 1127.18 |
| FD | 282 | 104 | 1609.41 |
| FD | 289 | 134 | 1526.98 |
| FD | 291 | 116 | 1421.81 |
| FD | 293 | 110 | 1777.52 |
| FD | 295 | 101 | 1156.39 |
| FD | 297 | 101 | 829.49 |
| FD | 301 | 113 | 1434.31 |
| FD | 302 | 103 | 1778.57 |
| FD | 303 | 110 | 986.43 |
| FD | 305 | 144 | 849.68 |
| FD | 310 | 100 | 1064.53 |
| FD | 318 | 164 | 929.78 |
| FD | 321 | 143 | 1125.23 |
| FD | 330 | 112 | 1039.94 |
| FD | 333 | 100 | 1169.08 |
| FD | 340 | 129 | 984.47 |
| FD | 341 | 139 | 1231.86 |
| FD | 342 | 177 | 1148.35 |
| FD | 343 | 103 | 939.79 |
| FD | 345 | 144 | 1399.33 |
| FD | 346 | 136 | 907.6 |
| FD | 347 | 100 | 1305 |
| FD | 352 | 147 | 1387.65 |
| FD | 369 | 136 | 1266.67 |
| FD | 373 | 111 | 1001 |
| FD | 389 | 144 | 1146.04 |
| FD | 391 | 111 | 1209.13 |
| FD | 401 | 139 | 1800.79 |
| FD | 402 | 100 | 2224.76 |
| FD | 406 | 178 | 633.25 |
| FD | 407 | 101 | 909.76 |
| FD | 411 | 110 | 2410.53 |
| FD* | 8* | 102 | 825.06 |
| FD* | 17 | 80 | 577.82 |
| CD | 58 | 9 | 713.57 |
| CD | 60 | 7 | 753.56 |
| CD | 61 | 6 | 1067.14 |
| CD | 63 | 4 | 1125.29 |
| CD | 65 | 5 | 1176.42 |
| CD | 67 | 9 | 1034.84 |
| CD | 68 | 10 | 969.64 |
| CD | 69 | 10 | 1867.19 |
| CD | 70 | 9 | 1045.26 |
| CD | 71 | 7 | 551.37 |
| CD | 72 | 2 | 1026.79 |
| CD | 73 | 3 | 1117 |
| CD | 74 | 5 | 1079.51 |
| CD | 75 | 7 | 1029.25 |
| CD | 76 | 10 | 456.5 |
| CD | 77 | 3 | 822.63 |
| CD | 78 | 5 | 757.04 |
| CD | 79 | 9 | 501.04 |
| CD | 80 | 6 | 514.87 |
| CD | 81 | 7 | 542.05 |
| CD | 82 | 5 | 471.94 |
| CD | 84 | 7 | 1553.71 |
| CD | 85 | 8 | 1429.82 |
| CD | 86 | 2 | 1184.06 |
| CD | 87 | 7 | 1157.13 |
| CD | 88 | 7 | 675.78 |
| CD | 89 | 3 | 732.62 |
| CD | 90 | 6 | 1818.44 |
| CD | 91 | 10 | 784.15 |
| CD | 95 | 2 | 780.4 |
| CD | 97 | 4 | 765.02 |
| CD | 98 | 10 | 627.72 |
| CD | 99 | 10 | 501.82 |
| CD | 100 | 10 | 649.8 |
| CD | 101 | 5 | 463.59 |
| CD | 102 | 8 | 418.31 |
| CD | 103 | 10 | 486.56 |
| CD | 104 | 5 | 567.11 |

| | | | |
|---|---|---|---|
| CD | 105 | 4 | 600.85 |
| CD | 106 | 8 | 590.3 |
| CD | 107 | 3 | 662.19 |
| CD | 127 | 5 | 1137.27 |
| CD | 132 | 5 | 1146.5 |
| CD | 144 | 9 | 1237.83 |
| CD | 146 | 2 | 1336.53 |
| CD | 148 | 8 | 1028.87 |
| CD | 157 | 7 | 1417.33 |
| CD | 166 | 2 | 865.39 |
| CD | 177 | 3 | 896.57 |
| CD | 183 | 8 | 1011.07 |
| CD | 184 | 3 | 1074.85 |
| CD | 193 | 6 | 1207.44 |
| CD | 196 | 3 | 1356.54 |
| CD | 201 | 3 | 1267.44 |
| CD | 220 | 9 | 1425.11 |
| CD | 222 | 4 | 1584.5 |
| CD | 228 | 8 | 1532.17 |
| CD | 234 | 9 | 1606.52 |
| CD | 241 | 2 | 734.54 |
| CD | 243 | 6 | 1383.55 |
| CD | 250 | 5 | 1413.03 |
| CD | 251 | 4 | 874.87 |
| CD | 259 | 4 | 873.74 |
| CD | 283 | 2 | 2170.12 |
| CD | 287 | 3 | 2382.11 |
| CD | 296 | 8 | 729.72 |
| CD | 299 | 8 | 995.52 |
| CD | 304 | 2 | 979.31 |
| CD | 308 | 3 | 1217.92 |
| CD | 309 | 4 | 904.62 |
| CD | 315 | 4 | 1011.33 |
| CD | 323 | 4 | 1280.06 |
| CD | 356 | 4 | 1229.8 |
| CD | 366 | 4 | 1114.64 |
| CD | 371 | 3 | 1071.62 |
| CD | 372 | 6 | 2685.15 |
| CD | 376 | 9 | 1041.21 |
| CD | 378 | 2 | 922.1 |
| CD | 387 | 2 | 1290.16 |
| CD | 398 | 8 | 1201.88 |
| CD | 408 | 8 | 1164.44 |
| CD | 410 | 3 | 910.63 |
| CD | 413 | 2 | 1939.22 |
| CD | 415 | 4 | 2331.81 |
| CD | 416 | 9 | 2257.19 |
| CD | 417 | 9 | 1059.48 |
| CD | 418 | 9 | 1000.44 |
| CD | 419 | 2 | 1105.3 |
| CD | 420 | 3 | 1416.51 |
| CD | 422 | 7 | 1620.38 |
| CD | 423 | 5 | 1456.79 |
| CD | 426 | 2 | 1280.99 |
| CD | 430 | 8 | 2539.58 |
| CD | 433 | 7 | 1623.37 |
| CD | 434 | 4 | 1624.36 |
| CD | 436 | 2 | 2241.62 |
| CD | 437 | 3 | 2060.62 |
| CD | 438 | 4 | 1418.54 |
| CD | 439 | 5 | 1660.14 |
| CD | 440 | 8 | 969.64 |
| CD | 442 | 9 | 1192.99 |
| CD | 444 | 2 | 1223.71 |
| CD | 445 | 8 | 1101.67 |
| * extended FD | | | |

**Supplemental Table 2. List of all detected mutations**

| Group | DonorID | NumberOfDonations | SequencingDepth | Gene_Name | CHROM | TYPE | VariantAlleleFrequency (VAF) | EpigeneticModifier | CorrelationPlot |
|---|---|---|---|---|---|---|---|---|---|
| FD | 2 | 144 | 965.39 | NA | NA | NA | NA | NA | NA |
| FD | 3 | 115 | 1248.82 | DNMT3A | chr2 | SNP | 0.0065923 | YES | DNMT3A |
| FD | 3 | 115 | 1248.82 | DNMT3A | chr2 | SNP | 0.0275362 | YES | DNMT3A |
| FD | 3 | 115 | 1248.82 | DNMT3A | chr2 | SNP | 0.0321285 | YES | DNMT3A |
| FD | 4 | 120 | 958.07 | NA | NA | NA | NA | NA | NA |
| FD | 7 | 105 | 684.22 | LRRC4 | chr7 | SNP | 0.030853 | NO | NA |
| FD* | 8 | 102 | 825.06 | CREBBP | chr16 | SNP | 0.0145278 | YES | OTHER |
| FD* | 8 | 102 | 825.06 | DNMT3A | chr2 | SNP | 0.0210526 | YES | DNMT3A |
| FD* | 8 | 102 | 825.06 | TET2 | chr4 | INDEL | 0.0288684 | YES | TET2 |
| FD | 9 | 133 | 664.41 | NA | NA | NA | NA | NA | NA |
| FD | 10 | 125 | 844.18 | NA | NA | NA | NA | NA | NA |
| FD | 11 | 112 | 805.95 | DNMT3A | chr2 | SNP | 0.0276596 | YES | DNMT3A |
| FD | 12 | 176 | 1016.42 | TET2 | chr4 | INDEL | 0.0113636 | YES | TET2 |
| FD* | 17 | 80 | 577.82 | DNMT3A | chr2 | INDEL | 0.0225873 | YES | DNMT3A |
| FD | 18 | 104 | 963.99 | DNMT3A | chr2 | SNP | 0.0164931 | YES | DNMT3A |
| FD | 19 | 101 | 2114.96 | NA | NA | NA | NA | NA | NA |
| FD | 20 | 136 | 853.85 | NA | NA | NA | NA | NA | NA |
| FD | 21 | 101 | 834.39 | NA | NA | NA | NA | NA | NA |
| FD | 22 | 110 | 1071.38 | NA | NA | NA | NA | NA | NA |
| FD | 23 | 126 | 633.68 | EP300 | chr22 | SNP | 0.0122449 | YES | OTHER |
| FD | 26 | 153 | 639.95 | DNMT3A | chr2 | INDEL | 0.0182927 | YES | DNMT3A |
| FD | 27 | 107 | 809.51 | DNMT1 | chr19 | SNP | 0.0094637 | YES | OTHER |
| FD | 27 | 107 | 809.51 | TET2 | chr4 | SNP | 0.0233645 | YES | TET2 |
| FD | 27 | 107 | 809.51 | DNMT3A | chr2 | SNP | 0.037225 | YES | DNMT3A |
| FD | 29 | 109 | 842.99 | NA | NA | NA | NA | NA | NA |
| FD | 30 | 110 | 715.83 | NA | NA | NA | NA | NA | NA |
| FD | 31 | 138 | 699.5 | DNMT3A | chr2 | SNP | 0.0151515 | YES | DNMT3A |
| FD | 31 | 138 | 699.5 | DNMT3A | chr2 | INDEL | 0.0751174 | YES | DNMT3A |
| FD | 33 | 105 | 431.14 | DNMT3A | chr2 | INDEL | 0.0204082 | YES | DNMT3A |
| FD | 34 | 126 | 698.1 | NA | NA | NA | NA | NA | NA |
| FD | 35 | 104 | 873.91 | DNMT3A | chr2 | SNP | 0.0063341 | YES | DNMT3A |
| FD | 35 | 104 | 873.91 | DNMT3A | chr2 | INDEL | 0.0099256 | YES | DNMT3A |
| FD | 35 | 104 | 873.91 | KMT2C | chr7 | SNP | 0.0192132 | YES | OTHER |
| FD | 36 | 101 | 769.25 | NA | NA | NA | NA | NA | NA |
| FD | 38 | 151 | 1451.24 | ASXL2 | chr2 | SNP | 0.005614 | YES | OTHER |
| FD | 38 | 151 | 1451.24 | HNRNPK | chr9 | SNP | 0.0106222 | NO | NA |
| FD | 40 | 154 | 1155.28 | NA | NA | NA | NA | NA | NA |
| FD | 41 | 167 | 626.71 | NA | NA | NA | NA | NA | NA |
| FD | 42 | 124 | 774.9 | DNMT3A | chr2 | SNP | 0.0140351 | YES | DNMT3A |
| FD | 42 | 124 | 774.9 | DNMT3A | chr2 | SNP | 0.091858 | YES | DNMT3A |
| FD | 44 | 146 | 855.66 | NA | NA | NA | NA | NA | NA |
| FD | 45 | 101 | 802.71 | TET2 | chr4 | INDEL | 0.0127389 | YES | TET2 |
| FD | 45 | 101 | 802.71 | CRLF2 | chrX | SNP | 0.3 | NO | NA |
| FD | 46 | 125 | 804 | NA | NA | NA | NA | NA | NA |
| FD | 48 | 100 | 884.29 | NA | NA | NA | NA | NA | NA |
| FD | 50 | 127 | 536.82 | NA | NA | NA | NA | NA | NA |
| FD | 51 | 122 | 751.87 | ELANE | chr19 | SNP | 0.0102389 | NO | NA |
| FD | 51 | 122 | 751.87 | DNMT3A | chr2 | INDEL | 0.060844 | YES | DNMT3A |
| FD | 53 | 101 | 692.95 | NA | NA | NA | NA | NA | NA |
| FD | 56 | 119 | 720.45 | DNMT3A | chr2 | INDEL | 0.179669 | YES | DNMT3A |
| FD | 57 | 122 | 1056.89 | DNMT3A | chr2 | SNP | 0.0093857 | YES | DNMT3A |
| FD | 66 | 102 | 1045.96 | DNMT3A | chr2 | SNP | 0.0265957 | YES | DNMT3A |
| FD | 94 | 161 | 1019.4 | TET2 | chr4 | SNP | 0.0131579 | YES | TET2 |
| FD | 116 | 102 | 1181.78 | TET2 | chr4 | SNP | 0.0140625 | YES | TET2 |
| FD | 122 | 107 | 1837.21 | DNMT3A | chr2 | SNP | 0.0096043 | YES | DNMT3A |
| FD | 123 | 115 | 1400.59 | DNMT3A | chr2 | SNP | 0.014549 | YES | DNMT3A |
| FD | 123 | 115 | 1400.59 | ASXL1 | chr20 | INDEL | 0.0460594 | YES | OTHER |
| FD | 124 | 128 | 1628.78 | SF3B1 | chr2 | SNP | 0.0102564 | NO | NA |
| FD | 124 | 128 | 1628.78 | DNMT3A | chr2 | SNP | 0.0112254 | YES | DNMT3A |
| FD | 126 | 130 | 1169.06 | NA | NA | NA | NA | NA | NA |
| FD | 136 | 111 | 1308.2 | DNMT3A | chr2 | SNP | 0.0082902 | YES | DNMT3A |
| FD | 143 | 134 | 1481.45 | KMT2C | chr7 | SNP | 0.04323 | YES | OTHER |
| FD | 147 | 140 | 901.08 | NA | NA | NA | NA | NA | NA |
| FD | 150 | 110 | 2003.89 | NA | NA | NA | NA | NA | NA |
| FD | 152 | 131 | 1473.09 | NA | NA | NA | NA | NA | NA |
| FD | 154 | 102 | 973 | BCR | chr22 | INDEL | 0.0397351 | NO | NA |
| FD | 167 | 101 | 1077 | NA | NA | NA | NA | NA | NA |
| FD | 190 | 163 | 1064.58 | NA | NA | NA | NA | NA | NA |
| FD | 200 | 111 | 1432.01 | DNMT3A | chr2 | SNP | 0.0215664 | YES | DNMT3A |
| FD | 208 | 137 | 1369.68 | NA | NA | NA | NA | NA | NA |
| FD | 224 | 126 | 1440.35 | NA | NA | NA | NA | NA | NA |
| FD | 231 | 101 | 1470.76 | NA | NA | NA | NA | NA | NA |
| FD | 239 | 126 | 1385.89 | NA | NA | NA | NA | NA | NA |
| FD | 244 | 121 | 2061.18 | BCR | chr22 | SNP | 0.0152513 | NO | NA |
| FD | 244 | 121 | 2061.18 | TET2 | chr4 | SNP | 0.0212187 | YES | TET2 |
| FD | 244 | 121 | 2061.18 | DNMT3A | chr2 | SNP | 0.1382037 | YES | DNMT3A |
| FD | 245 | 106 | 1501.18 | NA | NA | NA | NA | NA | NA |
| FD | 248 | 111 | 919.07 | DNMT3A | chr2 | INDEL | 0.0286807 | YES | DNMT3A |
| FD | 253 | 103 | 1277.43 | DNMT3A | chr2 | SNP | 0.007014 | YES | DNMT3A |
| FD | 256 | 103 | 1586.7 | CBL | chr11 | SNP | 0.0130152 | NO | NA |
| FD | 256 | 103 | 1586.7 | JAK3 | chr19 | SNP | 0.0635593 | NO | NA |
| FD | 257 | 125 | 1202.03 | DNMT3A | chr2 | SNP | 0.0064 | YES | DNMT3A |
| FD | 261 | 149 | 1070.68 | EED | chr11 | SNP | 0.0086207 | YES | OTHER |
| FD | 264 | 172 | 1048.1 | WAS | chrX | SNP | 0.01373 | NO | NA |
| FD | 264 | 172 | 1048.1 | KMT2C | chr7 | SNP | 0.021164 | YES | OTHER |
| FD | 266 | 140 | 1024.56 | NA | NA | NA | NA | NA | NA |
| FD | 268 | 132 | 1024.04 | NA | NA | NA | NA | NA | NA |
| FD | 269 | 109 | 1211.45 | CTCF | chr16 | SNP | 0.0189394 | NO | NA |
| FD | 269 | 109 | 1211.45 | KMT2C | chr7 | COMPLEX | 0.1280559 | YES | OTHER |
| FD | 270 | 100 | 1088.81 | TET2 | chr4 | SNP | 0.0088409 | YES | TET2 |
| FD | 270 | 100 | 1088.81 | DNMT3A | chr2 | INDEL | 0.0395683 | YES | DNMT3A |
| FD | 271 | 120 | 952.81 | NA | NA | NA | NA | NA | NA |
| FD | 273 | 135 | 1548.4 | DNMT3A | chr2 | SNP | 0.013459 | YES | DNMT3A |
| FD | 274 | 128 | 1061.47 | NA | NA | NA | NA | NA | NA |
| FD | 278 | 111 | 2289.06 | BRINP3 | chr1 | SNP | 0.0078377 | NO | NA |
| FD | 280 | 133 | 1127.18 | KMT2C | chr7 | SNP | 0.2608696 | YES | OTHER |
| FD | 282 | 104 | 1609.41 | KAT6A | chr8 | INDEL | 0.0124052 | YES | OTHER |
| FD | 289 | 134 | 1526.98 | KDM6A | chrX | SNP | 0.0225806 | YES | OTHER |

| | | | | | | | | | |
|---|---|---|---|---|---|---|---|---|---|
| FD | 291 | 116 | 1421.81 | TET2 | chr4 | SNP | 0.0212528 | YES | TET2 |
| FD | 291 | 116 | 1421.81 | DNMT3A | chr2 | SNP | 0.1338742 | YES | DNMT3A |
| FD | 293 | 110 | 1777.52 | NA | NA | NA | NA | NA | NA |
| FD | 295 | 101 | 1156.39 | NA | NA | NA | NA | NA | NA |
| FD | 297 | 101 | 829.49 | NA | NA | NA | NA | NA | NA |
| FD | 301 | 113 | 1434.31 | NA | NA | NA | NA | NA | NA |
| FD | 302 | 103 | 1778.57 | RUNX1 | chr21 | SNP | 0.0402685 | NO | NA |
| FD | 303 | 110 | 986.43 | TET2 | chr4 | SNP | 0.0107095 | YES | TET2 |
| FD | 303 | 110 | 986.43 | TET2 | chr4 | SNP | 0.0433071 | YES | TET2 |
| FD | 305 | 144 | 849.68 | NA | NA | NA | NA | NA | NA |
| FD | 310 | 100 | 1064.53 | DNMT3A | chr2 | SNP | 0.0069009 | YES | DNMT3A |
| FD | 318 | 164 | 929.78 | NA | NA | NA | NA | NA | NA |
| FD | 321 | 143 | 1125.23 | DNMT3A | chr2 | SNP | 0.0773196 | YES | DNMT3A |
| FD | 330 | 112 | 1039.94 | NA | NA | NA | NA | NA | NA |
| FD | 333 | 100 | 1169.08 | NA | NA | NA | NA | NA | NA |
| FD | 340 | 129 | 984.47 | NA | NA | NA | NA | NA | NA |
| FD | 341 | 139 | 1231.86 | KRAS | chr12 | SNP | 0.0091848 | NO | NA |
| FD | 341 | 139 | 1231.86 | SRSF2 | chr17 | SNP | 0.045045 | NO | NA |
| FD | 342 | 177 | 1148.35 | NA | NA | NA | NA | NA | NA |
| FD | 343 | 103 | 939.79 | DNMT3A | chr2 | SNP | 0.0067822 | YES | DNMT3A |
| FD | 345 | 144 | 1399.33 | NA | NA | NA | NA | NA | NA |
| FD | 346 | 136 | 907.6 | FAM154B | chr15 | SNP | 0.0338983 | NO | NA |
| FD | 347 | 100 | 1305 | TET2 | chr4 | SNP | 0.0086806 | YES | TET2 |
| FD | 347 | 100 | 1305 | TET2 | chr4 | INDEL | 0.009768 | YES | TET2 |
| FD | 347 | 100 | 1305 | MPL | chr1 | SNP | 0.0130719 | NO | NA |
| FD | 347 | 100 | 1305 | MPL | chr1 | SNP | 0.0132013 | NO | NA |
| FD | 347 | 100 | 1305 | MPL | chr1 | SNP | 0.013289 | NO | NA |
| FD | 352 | 147 | 1387.65 | SH2B3 | chr12 | SNP | 0.0154096 | NO | NA |
| FD | 352 | 147 | 1387.65 | SH2B3 | chr12 | INDEL | 0.0169753 | NO | NA |
| FD | 352 | 147 | 1387.65 | DNMT3A | chr2 | SNP | 0.0440613 | YES | DNMT3A |
| FD | 369 | 136 | 1266.67 | TET2 | chr4 | SNP | 0.073955 | YES | TET2 |
| FD | 373 | 111 | 1001 | TERT | chr5 | SNP | 0.0196078 | NO | NA |
| FD | 373 | 111 | 1001 | ATM | chr11 | INDEL | 0.0233236 | NO | NA |
| FD | 389 | 144 | 1146.04 | KRAS | chr12 | SNP | 0.0253353 | NO | NA |
| FD | 389 | 144 | 1146.04 | IKZF1 | chr7 | SNP | 0.0414938 | NO | NA |
| FD | 391 | 111 | 1209.13 | NA | NA | NA | NA | NA | NA |
| FD | 401 | 139 | 1800.79 | NA | NA | NA | NA | NA | NA |
| FD | 402 | 100 | 2224.76 | SMC1A | chrX | SNP | 0.0104948 | NO | NA |
| FD | 402 | 100 | 2224.76 | TET2 | chr4 | INDEL | 0.0145429 | YES | TET2 |
| FD | 406 | 178 | 633.25 | EED | chr11 | SNP | 0.0201613 | YES | OTHER |
| FD | 406 | 178 | 633.25 | STAT3 | chr17 | SNP | 0.0223048 | NO | NA |
| FD | 407 | 101 | 909.76 | NA | NA | NA | NA | NA | NA |
| FD | 411 | 110 | 2410.53 | SF3B1 | chr2 | SNP | 0.0152975 | NO | NA |
| CD | 58 | 9 | 713.57 | NA | NA | NA | NA | NA | NA |
| CD | 60 | 7 | 753.56 | NA | NA | NA | NA | NA | NA |
| CD | 61 | 6 | 1067.14 | NA | NA | NA | NA | NA | NA |
| CD | 63 | 4 | 1125.29 | DDX41 | chr5 | SNP | 0.0575658 | NO | NA |
| CD | 65 | 5 | 1176.42 | NA | NA | NA | NA | NA | NA |
| CD | 67 | 9 | 1034.84 | NA | NA | NA | NA | NA | NA |
| CD | 68 | 10 | 969.64 | NA | NA | NA | NA | NA | NA |
| CD | 69 | 10 | 1867.19 | HNRNPK | chr9 | INDEL | 0.0063839 | NO | NA |
| CD | 69 | 10 | 1867.19 | DNMT3A | chr2 | INDEL | 0.0077419 | YES | DNMT3A |
| CD | 70 | 9 | 1045.26 | TERT | chr5 | SNP | 0.0102421 | NO | NA |
| CD | 70 | 9 | 1045.26 | FIP1L1 | chr4 | SNP | 0.0267062 | NO | NA |
| CD | 71 | 7 | 551.37 | SMC1A | chrX | SNP | 0.1501976 | NO | NA |
| CD | 72 | 2 | 1026.79 | DNMT3A | chr2 | SNP | 0.0210526 | YES | DNMT3A |
| CD | 73 | 3 | 1117 | TET2 | chr4 | INDEL | 0.0113493 | YES | TET2 |
| CD | 74 | 5 | 1079.51 | NA | NA | NA | NA | NA | NA |
| CD | 75 | 7 | 1029.25 | DNMT3A | chr2 | INDEL | 0.0192593 | YES | DNMT3A |
| CD | 76 | 10 | 456.5 | NA | NA | NA | NA | NA | NA |
| CD | 77 | 3 | 822.63 | NA | NA | NA | NA | NA | NA |
| CD | 78 | 5 | 757.04 | TET2 | chr4 | SNP | 0.0343511 | YES | TET2 |
| CD | 78 | 5 | 757.04 | TET2 | chr4 | SNP | 0.0564706 | YES | TET2 |
| CD | 79 | 9 | 501.04 | ANKRD26 | chr10 | SNP | 0.0172414 | NO | NA |
| CD | 80 | 6 | 514.87 | NA | NA | NA | NA | NA | NA |
| CD | 81 | 7 | 542.05 | NA | NA | NA | NA | NA | NA |
| CD | 82 | 5 | 471.94 | NA | NA | NA | NA | NA | NA |
| CD | 84 | 7 | 1553.71 | NA | NA | NA | NA | NA | NA |
| CD | 85 | 8 | 1429.82 | NOTCH1 | chr9 | SNP | 0.0069174 | NO | NA |
| CD | 85 | 8 | 1429.82 | FAM47A | chrX | SNP | 0.0163551 | NO | NA |
| CD | 85 | 8 | 1429.82 | JAK3 | chr19 | SNP | 0.0318991 | NO | NA |
| CD | 86 | 2 | 1184.06 | NA | NA | NA | NA | NA | NA |
| CD | 87 | 7 | 1157.13 | RUNX1 | chr21 | SNP | 0.0106257 | NO | NA |
| CD | 88 | 7 | 675.78 | NA | NA | NA | NA | NA | NA |
| CD | 89 | 3 | 732.62 | NA | NA | NA | NA | NA | NA |
| CD | 90 | 6 | 1818.44 | NA | NA | NA | NA | NA | NA |
| CD | 91 | 10 | 784.15 | NA | NA | NA | NA | NA | NA |
| CD | 95 | 2 | 780.4 | NA | NA | NA | NA | NA | NA |
| CD | 97 | 4 | 765.02 | NA | NA | NA | NA | NA | NA |
| CD | 98 | 10 | 627.72 | BCR | chr22 | SNP | 0.0296736 | NO | NA |
| CD | 99 | 10 | 501.82 | NA | NA | NA | NA | NA | NA |
| CD | 100 | 10 | 649.8 | NA | NA | NA | NA | NA | NA |
| CD | 101 | 5 | 463.59 | NA | NA | NA | NA | NA | NA |
| CD | 102 | 8 | 418.31 | FAM47A | chrX | SNP | 0.0431655 | NO | NA |
| CD | 103 | 10 | 486.56 | NA | NA | NA | NA | NA | NA |
| CD | 104 | 5 | 567.11 | NA | NA | NA | NA | NA | NA |
| CD | 105 | 4 | 600.85 | NA | NA | NA | NA | NA | NA |
| CD | 106 | 8 | 590.3 | U2AF1 | chr21 | SNP | 0.0263975 | NO | NA |
| CD | 106 | 8 | 590.3 | ASXL1 | chr20 | INDEL | 0.030445 | YES | OTHER |
| CD | 107 | 3 | 662.19 | ATM | chr11 | SNP | 0.0116822 | NO | NA |
| CD | 127 | 5 | 1137.27 | DNMT3A | chr2 | SNP | 0.0113924 | YES | DNMT3A |
| CD | 132 | 5 | 1146.5 | TET2 | chr4 | SNP | 0.0248619 | YES | TET2 |
| CD | 132 | 5 | 1146.5 | WAS | chrX | SNP | 0.0677966 | NO | NA |
| CD | 132 | 5 | 1146.5 | DNMT3A | chr2 | SNP | 0.3051085 | YES | DNMT3A |
| CD | 144 | 9 | 1237.83 | NA | NA | NA | NA | NA | NA |
| CD | 146 | 2 | 1336.53 | DNMT3A | chr2 | SNP | 0.01375 | YES | DNMT3A |
| CD | 148 | 8 | 1028.87 | NA | NA | NA | NA | NA | NA |
| CD | 157 | 7 | 1417.33 | NA | NA | NA | NA | NA | NA |
| CD | 166 | 2 | 865.39 | NA | NA | NA | NA | NA | NA |

**Supplemental Table 2. List of all detected mutations**

| | | | | | | | | | |
|---|---|---|---|---|---|---|---|---|---|
| CD | 177 | 3 | 896.57 | ASXL2 | chr2 | INDEL | 0.0145228 | YES | OTHER |
| CD | 177 | 3 | 896.57 | DNMT3A | chr2 | SNP | 0.0155521 | YES | DNMT3A |
| CD | 177 | 3 | 896.57 | DNMT3A | chr2 | INDEL | 0.0636833 | YES | DNMT3A |
| CD | 183 | 8 | 1011.07 | BRCA2 | chr13 | SNP | 0.0162866 | NO | NA |
| CD | 183 | 8 | 1011.07 | DNMT3A | chr2 | SNP | 0.0193322 | YES | DNMT3A |
| CD | 184 | 3 | 1074.85 | NA | NA | NA | NA | NA | NA |
| CD | 193 | 6 | 1207.44 | SETBP1 | chr18 | SNP | 0.0181818 | YES | OTHER |
| CD | 193 | 6 | 1207.44 | DNMT3A | chr2 | INDEL | 0.0211679 | YES | DNMT3A |
| CD | 193 | 6 | 1207.44 | TET2 | chr4 | SNP | 0.0297806 | YES | TET2 |
| CD | 196 | 3 | 1356.54 | SF3B1 | chr2 | INDEL | 0.0108254 | NO | NA |
| CD | 201 | 3 | 1267.44 | DNM2 | chr19 | SNP | 0.04209 | NO | NA |
| CD | 220 | 9 | 1425.11 | NA | NA | NA | NA | NA | NA |
| CD | 222 | 4 | 1584.5 | CBL | chr11 | SNP | 0.0080175 | NO | NA |
| CD | 228 | 8 | 1532.17 | NA | NA | NA | NA | NA | NA |
| CD | 234 | 9 | 1606.52 | NA | NA | NA | NA | NA | NA |
| CD | 241 | 2 | 734.54 | NA | NA | NA | NA | NA | NA |
| CD | 243 | 6 | 1383.55 | NA | NA | NA | NA | NA | NA |
| CD | 250 | 5 | 1413.03 | NA | NA | NA | NA | NA | NA |
| CD | 251 | 4 | 874.87 | SH2B3 | chr12 | SNP | 0.0094439 | NO | NA |
| CD | 259 | 4 | 873.74 | NA | NA | NA | NA | NA | NA |
| CD | 283 | 2 | 2170.12 | HNRNPK | chr9 | INDEL | 0.0059752 | NO | NA |
| CD | 283 | 2 | 2170.12 | DNMT3A | chr2 | SNP | 0.0234443 | YES | DNMT3A |
| CD | 283 | 2 | 2170.12 | DNMT3A | chr2 | INDEL | 0.0244845 | YES | DNMT3A |
| CD | 283 | 2 | 2170.12 | NPAT | chr11 | INDEL | 0.352575 | NO | NA |
| CD | 287 | 3 | 2382.11 | NA | NA | NA | NA | NA | NA |
| CD | 296 | 8 | 729.72 | NA | NA | NA | NA | NA | NA |
| CD | 299 | 8 | 995.52 | NA | NA | NA | NA | NA | NA |
| CD | 304 | 2 | 979.31 | NA | NA | NA | NA | NA | NA |
| CD | 308 | 3 | 1217.92 | NA | NA | NA | NA | NA | NA |
| CD | 309 | 4 | 904.62 | NA | NA | NA | NA | NA | NA |
| CD | 315 | 4 | 1011.33 | NA | NA | NA | NA | NA | NA |
| CD | 323 | 4 | 1280.06 | NA | NA | NA | NA | NA | NA |
| CD | 356 | 4 | 1229.8 | BRCA2 | chr13 | SNP | 0.0465116 | NO | NA |
| CD | 366 | 4 | 1114.64 | NA | NA | NA | NA | NA | NA |
| CD | 371 | 3 | 1071.62 | DNMT3A | chr2 | SNP | 0.0131805 | YES | DNMT3A |
| CD | 371 | 3 | 1071.62 | DNM2 | chr19 | SNP | 0.0136986 | NO | NA |
| CD | 372 | 6 | 2685.15 | TET2 | chr4 | INDEL | 0.0062684 | YES | TET2 |
| CD | 372 | 6 | 2685.15 | DNMT3A | chr2 | SNP | 0.0091463 | YES | DNMT3A |
| CD | 376 | 9 | 1041.21 | KMT2C | chr7 | SNP | 0.048913 | YES | OTHER |
| CD | 378 | 2 | 922.1 | NA | NA | NA | NA | NA | NA |
| CD | 387 | 2 | 1290.16 | CUX1 | chr7 | SNP | 0.0168498 | NO | NA |
| CD | 398 | 8 | 1201.88 | NA | NA | NA | NA | NA | NA |
| CD | 408 | 8 | 1164.44 | NA | NA | NA | NA | NA | NA |
| CD | 410 | 3 | 910.63 | IL7R | chr5 | SNP | 0.3724792 | NO | NA |
| CD | 413 | 2 | 1939.22 | NA | NA | NA | NA | NA | NA |
| CD | 415 | 4 | 2331.81 | TET2 | chr4 | SNP | 0.0263415 | YES | TET2 |
| CD | 416 | 9 | 2257.19 | GJB3 | chr1 | SNP | 0.0051787 | NO | NA |
| CD | 416 | 9 | 2257.19 | TAL1 | chr1 | SNP | 0.0066815 | NO | NA |
| CD | 417 | 9 | 1059.48 | NA | NA | NA | NA | NA | NA |
| CD | 418 | 9 | 1000.44 | NA | NA | NA | NA | NA | NA |
| CD | 419 | 2 | 1105.3 | NA | NA | NA | NA | NA | NA |
| CD | 420 | 3 | 1416.51 | DNMT3A | chr2 | SNP | 0.0151362 | YES | DNMT3A |
| CD | 420 | 3 | 1416.51 | DNMT3A | chr2 | SNP | 0.037037 | YES | DNMT3A |
| CD | 422 | 7 | 1620.38 | BRCA1 | chr17 | SNP | 0.0082353 | NO | NA |
| CD | 422 | 7 | 1620.38 | ABL1 | chr9 | SNP | 0.0084122 | NO | NA |
| CD | 423 | 5 | 1456.79 | DNMT3A | chr2 | INDEL | 0.064877 | YES | DNMT3A |
| CD | 426 | 2 | 1280.99 | NA | NA | NA | NA | NA | NA |
| CD | 430 | 8 | 2539.58 | CHEK2 | chr22 | INDEL | 0.0062406 | NO | NA |
| CD | 430 | 8 | 2539.58 | DNMT3A | chr2 | SNP | 0.0069212 | YES | DNMT3A |
| CD | 433 | 7 | 1623.37 | DNMT3A | chr2 | SNP | 0.0178069 | YES | DNMT3A |
| CD | 434 | 4 | 1624.36 | DNMT3A | chr2 | SNP | 0.012973 | YES | DNMT3A |
| CD | 434 | 4 | 1624.36 | DNMT3A | chr2 | SNP | 0.0143296 | YES | DNMT3A |
| CD | 434 | 4 | 1624.36 | DNMT3A | chr2 | INDEL | 0.0257787 | YES | DNMT3A |
| CD | 434 | 4 | 1624.36 | SMC1A | chrX | SNP | 0.0388889 | NO | NA |
| CD | 436 | 2 | 2241.62 | NA | NA | NA | NA | NA | NA |
| CD | 437 | 3 | 2060.62 | NA | NA | NA | NA | NA | NA |
| CD | 438 | 4 | 1418.54 | NA | NA | NA | NA | NA | NA |
| CD | 439 | 5 | 1660.14 | NA | NA | NA | NA | NA | NA |
| CD | 440 | 8 | 969.64 | NA | NA | NA | NA | NA | NA |
| CD | 442 | 9 | 1192.99 | NA | NA | NA | NA | NA | NA |
| CD | 444 | 2 | 1223.71 | DNMT3A | chr2 | INDEL | 0.0077419 | YES | DNMT3A |
| CD | 445 | 8 | 1101.67 | NA | NA | NA | NA | NA | NA |
| * extended FD | | | | | | | | | |

**Supplemental Table 3. List of all DNMT3A mutations**

| Group | UMIDepth | DonorID | NumberOfDonations | CHROM | POS | ID | REF | ALT | QUAL | FILTER | TYPE | DP | UMT | VMT | VMF | Allele | Annotation | Annotation_Impact | Gene_Name | HGVS.c | Exchange | IDandPos | Stability Score | Premature STOP? |
|---|---|---|---|---|---|---|---|---|---|---|---|---|---|---|---|---|---|---|---|---|---|---|---|---|
| CD | 1029.25 | 75 | 7 | chr2 | 25469990 | . | C | CA | 12 | PASS | INDEL | 5861 | 675 | 13 | 0.0192593 | CA | frameshift_variant | HIGH | DNMT3A | c.1051dupT | p.Cys351fs | DNMT3A c.1051dupT | NA | STOP |
| CD | 1623.37 | 433 | 7 | chr2 | 25469919 | . | C | T | 15 | PASS | SNP | 5423 | 1067 | 19 | 0.0178069 | T | splice_donor_variant&intron_variant | HIGH | DNMT3A | c.1122+1G>A | NA | DNMT3A c.1122+1G>A | NA | NOT STOP |
| CD | 1207.44 | 193 | 6 | chr2 | 25469588 | . | CA | C | 24 | PASS | INDEL | 8936 | 1370 | 29 | 0.0211679 | C | frameshift_variant | HIGH | DNMT3A | c.1179delT | p.Ser393fs | DNMT3A c.1179delT | NA | STOP |
| CD | 1416.51 | 420 | 3 | chr2 | 25467023 | . | C | T | 38 | PASS | SNP | 4067 | 1080 | 40 | 0.037037 | T | splice_donor_variant&intron_variant | HIGH | DNMT3A | c.1851+1G>A | NA | DNMT3A c.1851+1G>A | NA | NOT STOP |
| CD | 2170.12 | 283 | 2 | chr2 | 25466784 | . | AAG | A | 67 | PASS | INDEL | 14465 | 3880 | 95 | 0.0244845 | A | frameshift_variant | HIGH | DNMT3A | c.1917_1918delCT | p.Phe640fs | DNMT3A c.1917_1918delCT | NA | STOP |
| CD | 1416.51 | 420 | 3 | chr2 | 25464456 | . | T | C | 11 | PASS | SNP | 4131 | 991 | 15 | 0.0151362 | C | missense_variant | MODERATE | DNMT3A | c.2057A>G | p.Asp686Gly | DNMT3A c.2057A>G | NA | NOT STOP |
| CD | 1624.36 | 434 | 4 | chr2 | 25463576 | . | A | C | 9 | PASS | SNP | 5430 | 925 | 12 | 0.012973 | C | structural_interaction_variant | HIGH | DNMT3A | c.2106T>G | p.Asp702Glu | DNMT3A c.2106T>G | NA | NOT STOP |
| CD | 1026.79 | 72 | 2 | chr2 | 25463568 | COSM1583102 | A | G | 11 | PASS | SNP | 3991 | 570 | 12 | 0.0210526 | G | missense_variant | MODERATE | DNMT3A | c.2114T>C | p.Ile705Thr | DNMT3A c.2114T>C | 0,3875 | NOT STOP |
| CD | 896.57 | 177 | 3 | chr2 | 25463549 | . | AT | A | 46 | PASS | INDEL | 4942 | 581 | 37 | 0.0636833 | A | frameshift_variant | HIGH | DNMT3A | c.2132delA | p.Asn711fs | DNMT3A c.2132delA | NA | STOP |
| CD | 1867.19 | 69 | 10 | chr2 | 25463291 | . | GAACTCAAA | G | 6 | PASS | INDEL | 5422 | 1550 | 12 | 0.0077419 | G | frameshift_variant | HIGH | DNMT3A | c.2194_2201delTTTGAGTT | p.Phe732fs | DNMT3A c.2194_2201delTTTGAGTT | NA | STOP |
| CD | 1137.27 | 127 | 5 | chr2 | 25463289 | COSM133126 | T | C | 7 | PASS | SNP | 5360 | 790 | 9 | 0.0113924 | C | missense_variant | MODERATE | DNMT3A | c.2204A>G | p.Tyr735Cys | DNMT3A c.2204A>G | 0,8631 | NOT STOP |
| CD | 1223.71 | 444 | 2 | chr2 | 25463235 | COSM133723 | CAGA | C | 7 | PASS | INDEL | 14479 | 1550 | 12 | 0.0077419 | C | protein_protein_contact | HIGH | DNMT3A | c.2255_2257delTCT | p.Phe752fs | DNMT3A c.2255_2257delTCT | NA | |
| CD | 1624.36 | 434 | 4 | chr2 | 25463235 | COSM133723 | CAGA | C | 36 | PASS | INDEL | 10019 | 1862 | 48 | 0.0257787 | C | structural_interaction_variant | HIGH | DNMT3A | c.2255_2257delTCT | p.Phe752fs | DNMT3A c.2255_2257delTCT | NA | |
| CD | 2170.12 | 283 | 2 | chr2 | 25463232 | . | A | T | 56 | PASS | SNP | 13133 | 3455 | 81 | 0.0234443 | T | missense_variant | MODERATE | DNMT3A | c.2261T>A | p.Leu754His | DNMT3A c.2261T>A | NA | NOT STOP |
| CD | 2539.58 | 430 | 8 | chr2 | 25463212 | . | T | C | 11 | PASS | SNP | 12828 | 4479 | 31 | 0.0069212 | C | missense_variant | MODERATE | DNMT3A | c.2281A>G | p.Met761Val | DNMT3A c.2281A>G | NA | NOT STOP |
| CD | 1336.53 | 146 | 2 | chr2 | 25462068 | COSM1583121 | A | G | 9 | PASS | SNP | 4647 | 800 | 11 | 0.01375 | G | missense_variant | MODERATE | DNMT3A | c.2339T>C | p.Ile780Thr | DNMT3A c.2339T>C | 0,4926 | NOT STOP |
| CD | 2685.15 | 372 | 6 | chr2 | 25459806 | COSM1583124 | T | C | 8 | PASS | SNP | 5301 | 1640 | 15 | 0.0091463 | C | missense_variant&splice_region_variant | MODERATE | DNMT3A | c.2477A>G | p.Lys826Arg | DNMT3A c.2477A>G | 1,0628 | NOT STOP |
| CD | 896.57 | 177 | 3 | chr2 | 25458652 | . | T | C | 9 | PASS | SNP | 5143 | 643 | 10 | 0.0155521 | C | missense_variant | MODERATE | DNMT3A | c.2521A>G | p.Lys841Glu | DNMT3A c.2521A>G | NA | NOT STOP |
| CD | 1456.79 | 423 | 5 | chr2 | 25458596 | . | TA | T | 69 | PASS | INDEL | 5259 | 894 | 58 | 0.064877 | T | frameshift_variant | HIGH | DNMT3A | c.2576delT | p.Leu859fs | DNMT3A c.2576delT | NA | STOP |
| CD | 1011.07 | 183 | 8 | chr2 | 25458595 | COSM231568 | A | G | 10 | PASS | SNP | 4259 | 569 | 11 | 0.0193322 | G | missense_variant | MODERATE | DNMT3A | c.2578T>C | p.Trp860Arg | DNMT3A c.2578T>C | 0,9209 | NOT STOP |
| CD | 1624.36 | 434 | 4 | chr2 | 25458595 | COSM231568 | A | G | 10 | PASS | SNP | 6675 | 977 | 14 | 0.0143296 | G | missense_variant | MODERATE | DNMT3A | c.2578T>C | p.Trp860Arg | DNMT3A c.2578T>C | 0,9209 | NOT STOP |
| CD | 1071.62 | 371 | 3 | chr2 | 25457243 | COSM1166704;COSM53042 | G | A | 14 | PASS | SNP | 8986 | 1745 | 23 | 0.0131805 | A | missense_variant | MODERATE | DNMT3A | c.2644C>T | p.Arg882Cys | DNMT3A c.2644C>T | NA | NOT STOP |
| CD | 1146.5 | 132 | 5 | chr2 | 25457242 | COSM442676;COSM52944 | C | T | 200 | PASS | SNP | 9591 | 1429 | 436 | 0.3051085 | T | missense_variant | MODERATE | DNMT3A | c.2645G>A | p.Arg882His | DNMT3A c.2645G>A | 0,9765 | NOT STOP |
| FD | 1088.81 | 270 | 100 | chr2 | 25470472 | COSM133727 | GC | G | 34 | PASS | INDEL | 7848 | 834 | 33 | 0.0395683 | G | frameshift_variant | HIGH | DNMT3A | c.1001delG | p.Gly334fs | DNMT3A c.1001delG | NA | STOP |
| FD | 774.9 | 42 | 124 | chr2 | 25470002 | . | A | G | 7 | PASS | SNP | 6174 | 570 | 8 | 0.0140351 | G | missense_variant | MODERATE | DNMT3A | c.1040T>C | p.Leu347Pro | DNMT3A c.1040T>C | NA | NOT STOP |
| FD | 1421.81 | 291 | 116 | chr2 | 25469945 | . | C | T | 190 | PASS | SNP | 7838 | 986 | 132 | 0.1338742 | T | missense_variant | MODERATE | DNMT3A | c.1097G>A | p.Arg366His | DNMT3A c.1097G>A | 0,7082 | NOT STOP |
| FD | 431.14 | 33 | 105 | chr2 | 25469613 | COSM1717669;COSM1717670 | CG | C | 6 | PASS | INDEL | 3562 | 245 | 5 | 0.0204082 | C | frameshift_variant | HIGH | DNMT3A | c.1154delC | p.Pro385fs | DNMT3A c.1154delC | NA | STOP |
| FD | 639.95 | 26 | 153 | chr2 | 25469083 | . | TC | T | 11 | PASS | INDEL | 4805 | 656 | 12 | 0.0182927 | T | frameshift_variant | HIGH | DNMT3A | c.1374delG | p.Lys459fs | DNMT3A c.1374delG | NA | STOP |
| FD | 919.07 | 248 | 111 | chr2 | 25468928 | . | GC | G | 16 | PASS | INDEL | 4400 | 523 | 15 | 0.0286807 | G | frameshift_variant | HIGH | DNMT3A | c.1434delG | p.Leu479fs | DNMT3A c.1434delG | NA | STOP |
| FD | 1202.03 | 257 | 125 | chr2 | 25467467 | . | A | T | 6 | PASS | SNP | 11472 | 1875 | 12 | 0.0064 | T | missense_variant | MODERATE | DNMT3A | c.1609T>A | p.Cys537Ser | DNMT3A c.1609T>A | 0,4449 | NOT STOP |
| FD | 1432.01 | 200 | 111 | chr2 | 25467408 | . | C | T | 16 | PASS | SNP | 3563 | 881 | 19 | 0.0215664 | T | splice_donor_variant&intron_variant | HIGH | DNMT3A | c.1667+1G>A | NA | DNMT3A c.1667+1G>A | NA | NOT STOP |
| FD | 873.91 | 35 | 104 | chr2 | 25467104 | . | T | TGC | 6 | PASS | INDEL | 5687 | 806 | 8 | 0.0099256 | TGC | frameshift_variant | HIGH | DNMT3A | c.1770_1771insGC | p.Thr591fs | DNMT3A c.1770_1771insGC | NA | STOP |
| FD | 1248.82 | 3 | 115 | chr2 | 25466800 | COSM1407108;COSM87012 | G | A | 6 | PASS | SNP | 9796 | 1972 | 13 | 0.0065923 | A | structural_interaction_variant | HIGH | DNMT3A | c.1903C>T | p.Arg635Trp | DNMT3A c.1903C>T | 0,4060 | NOT STOP |
| FD | 1308.2 | 136 | 111 | chr2 | 25466800 | COSM1407108;COSM87012 | G | A | 9 | PASS | SNP | 16504 | 1930 | 16 | 0.0082902 | A | structural_interaction_variant | HIGH | DNMT3A | c.1903C>T | p.Arg635Trp | DNMT3A c.1903C>T | 0,4060 | NOT STOP |
| FD | 873.91 | 35 | 104 | chr2 | 25464525 | . | G | C | 6 | PASS | SNP | 8606 | 1263 | 8 | 0.0063341 | C | missense_variant | MODERATE | DNMT3A | c.1988C>G | p.Ser663Trp | DNMT3A c.1988C>G | 0,4803 | NOT STOP |
| FD | 751.87 | 51 | 122 | chr2 | 25464524 | . | CG | C | 73 | PASS | INDEL | 8683 | 1019 | 62 | 0.060844 | C | frameshift_variant | HIGH | DNMT3A | c.1988delC | p.Ser663fs | DNMT3A c.1988delC | NA | STOP |
| FD | 1628.78 | 124 | 128 | chr2 | 25463600 | . | C | T | 8 | PASS | SNP | 4293 | 1069 | 12 | 0.0112254 | T | splice_acceptor_variant&intron_variant | HIGH | DNMT3A | c.2083-1G>A | NA | DNMT3A c.2083-1G>A | NA | NOT STOP |
| FD | 699.5 | 31 | 138 | chr2 | 25463297 | . | AAAG | A | 42 | PASS | INDEL | 4982 | 426 | 32 | 0.0751174 | A | disruptive_inframe_deletion | MODERATE | DNMT3A | c.2193_2195delCTT | p.Phe732del | DNMT3A c.2193_2195delCTT | NA | |
| FD | 774.9 | 42 | 124 | chr2 | 25463296 | . | C | A | 71 | PASS | SNP | 3927 | 479 | 44 | 0.091858 | A | stop_gained | HIGH | DNMT3A | c.2197G>T | p.Glu733* | DNMT3A c.2197G>T | NA | STOP |
| FD | 1387.65 | 352 | 147 | chr2 | 25463292 | . | A | G | 48 | PASS | SNP | 5810 | 1044 | 46 | 0.0440613 | G | missense_variant | MODERATE | DNMT3A | c.2201T>C | p.Phe734Ser | DNMT3A c.2201T>C | NA | NOT STOP |
| FD | 963.99 | 18 | 104 | chr2 | 25463287 | COSM231560 | G | A | 15 | PASS | SNP | 8939 | 1152 | 19 | 0.0164931 | A | protein_protein_contact | HIGH | DNMT3A | c.2206C>T | p.Arg736Cys | DNMT3A c.2206C>T | 0,3159 | NOT STOP |
| FD | 1064.53 | 310 | 100 | chr2 | 25463248 | COSM219133 | G | A | 6 | PASS | SNP | 8828 | 1594 | 11 | 0.0069009 | A | structural_interaction_variant | HIGH | DNMT3A | c.2245C>T | p.Arg749Cys | DNMT3A c.2245C>T | 0,3393 | NOT STOP |
| FD | 1056.89 | 57 | 122 | chr2 | 25463229 | . | A | G | 7 | PASS | SNP | 9377 | 1172 | 11 | 0.0093857 | G | missense_variant | MODERATE | DNMT3A | c.2264T>C | p.Phe755Ser | DNMT3A c.2264T>C | NA | NOT STOP |
| FD | 1125.23 | 321 | 143 | chr2 | 25463184 | COSM231549 | G | A | 75 | PASS | SNP | 4663 | 776 | 60 | 0.0773196 | A | missense_variant | MODERATE | DNMT3A | c.2309C>T | p.Ser770Leu | DNMT3A c.2309C>T | 0,4189 | NOT STOP |
| FD | 1248.82 | 3 | 115 | chr2 | 25463182 | COSM231563 | G | A | 18 | PASS | SNP | 4148 | 690 | 19 | 0.0275362 | A | stop_gained | HIGH | DNMT3A | c.2311C>T | p.Arg771* | DNMT3A c.2311C>T | NA | STOP |
| FD | 1548.4 | 273 | 135 | chr2 | 25463182 | COSM231563 | G | A | 8 | PASS | SNP | 5110 | 743 | 10 | 0.013459 | A | stop_gained | HIGH | DNMT3A | c.2311C>T | p.Arg771* | DNMT3A c.2311C>T | NA | STOP |
| FD | 805.95 | 11 | 112 | chr2 | 25462085 | . | C | A | 17 | PASS | SNP | 3420 | 470 | 13 | 0.0276596 | A | splice_acceptor_variant&intron_variant | HIGH | DNMT3A | c.2323-1G>T | NA | DNMT3A c.2323-1G>T | NA | NOT STOP |
| FD | 720.45 | 56 | 119 | chr2 | 25459847 | . | CT | C | 125 | PASS | INDEL | 3922 | 423 | 76 | 0.179669 | C | frameshift_variant | HIGH | DNMT3A | c.2435delA | p.Lys812fs | DNMT3A c.2435delA | NA | STOP |
| FD | 699.5 | 31 | 138 | chr2 | 25458661 | COSM231575 | T | C | 7 | PASS | SNP | 5729 | 528 | 8 | 0.0151515 | C | missense_variant | MODERATE | DNMT3A | c.2512A>G | p.Asn838Asp | DNMT3A c.2512A>G | 0,9860 | NOT STOP |
| FD | 1045.96 | 66 | 102 | chr2 | 25457285 | . | A | G | 15 | PASS | SNP | 7376 | 564 | 15 | 0.0265957 | G | protein_protein_contact | HIGH | DNMT3A | c.2602T>C | p.Phe868leu | DNMT3A c.2602T>C | NA | NOT STOP |
| FD | 1277.43 | 253 | 103 | chr2 | 25457243 | COSM1166704;COSM53042 | G | A | 7 | PASS | SNP | 11726 | 1996 | 14 | 0.007014 | A | missense_variant | MODERATE | DNMT3A | c.2644C>T | p.Arg882Cys | DNMT3A c.2644C>T | NA | NOT STOP |
| FD | 1837.21 | 122 | 107 | chr2 | 25457243 | COSM1166704;COSM53042 | G | A | 13 | PASS | SNP | 11184 | 2603 | 25 | 0.0096043 | A | missense_variant | MODERATE | DNMT3A | c.2644C>T | p.Arg882Cys | DNMT3A c.2644C>T | NA | NOT STOP |
| FD | 2061.18 | 244 | 121 | chr2 | 25457242 | COSM442676;COSM52944 | C | T | 200 | PASS | SNP | 13037 | 3162 | 437 | 0.1382037 | T | missense_variant | MODERATE | DNMT3A | c.2645G>A | p.Arg882His | DNMT3A c.2645G>A | 0,9765 | NOT STOP |
| FD | 939.79 | 343 | 103 | chr2 | 25457204 | . | C | G | 7 | PASS | SNP | 10791 | 1327 | 9 | 0.0067822 | G | structural_interaction_variant | HIGH | DNMT3A | c.2683G>C | p.Val895Leu | DNMT3A c.2683G>C | NA | NOT STOP |
| FD* | 577.82 | 17 | 80 | chr2 | 25470903 | . | CA | C | 11 | PASS | INDEL | 4478 | 487 | 11 | 0.0225873 | C | splice_donor_variant&intron_variant | HIGH | DNMT3A | c.855+2delT | NA | DNMT3A c.855+2delT | NA | NOT STOP |
| FD* | 825.06 | 8 | 102 | chr2 | 25470560 | COSM1169636 | C | T | 13 | PASS | SNP | 4890 | 665 | 14 | 0.0210526 | T | stop_gained | HIGH | DNMT3A | c.914G>A | p.Trp305* | DNMT3A c.914G>A | NA | STOP |
| FD | 1248.82 | 3 | 115 | chr2 | 25470559 | . | C | T | 37 | PASS | SNP | 5874 | 1245 | 40 | 0.0321285 | T | stop_gained | HIGH | DNMT3A | c.915G>A | p.Trp305* | DNMT3A c.915G>A | NA | STOP |
| FD | 809.51 | 27 | 107 | chr2 | 25470498 | . | G | C | 29 | PASS | SNP | 5058 | 591 | 22 | 0.037225 | C | missense_variant | MODERATE | DNMT3A | c.976C>G | p.Arg326Gly | DNMT3A c.976C>G | NA | NOT STOP |
| FD | 1400.59 | 123 | 115 | chr2 | 25470481 | . | G | T | 14 | PASS | SNP | 7617 | 1031 | 15 | 0.014549 | T | missense_variant | MODERATE | DNMT3A | c.993C>A | p.Phe331Leu | DNMT3A c.993C>A | NA | NOT STOP |

* extended FD

**Supplemental Table 4. List of all *TET2* mutations**

| Group | UMIDepth | VariantsTotal | DonorID | NumberOfDonations | CHROM | POS | ID | REF | ALT | QUAL | FILTER | TYPE | DP | UMT | VMT | VMF | Allele | Annotation | Annotation_Impact | Gene_Name | HGVS.c | Exchange | IDandPos |
|---|---|---|---|---|---|---|---|---|---|---|---|---|---|---|---|---|---|---|---|---|---|---|---|
| CD | 757.04 | 149 | 78 | 5 | chr4 | 106196309 | COSM51209 | C | T | 19 | PASS | SNP | 6420 | 524 | 18 | 0.0343511 | T | stop_gained | HIGH | TET2 | c.4705C>T | p.Gln1569* | TET2 c.4705C>T |
| CD | 757.04 | 149 | 78 | 5 | chr4 | 106197285 | COSM41741 | T | C | 30 | PASS | SNP | 3903 | 425 | 24 | 0.0564706 | C | missense_variant | MODERATE | TET2 | c.5681T>C | p.Ile1894Thr | TET2 c.5681T>C |
| FD | 802.71 | 164 | 45 | 101 | chr4 | 106157515 | . | CT | C | 8 | PASS | INDEL | 6188 | 785 | 10 | 0.0127389 | C | frameshift_variant | HIGH | TET2 | c.2480delT | p.Leu827fs | TET2 c.2480delT |
| FD | 809.51 | 193 | 27 | 107 | chr4 | 106162586 | . | G | A | 14 | PASS | SNP | 5627 | 642 | 15 | 0.0233645 | A | missense_variant&splice_region_variant | MODERATE | TET2 | c.3563G>A | p.Arg1188Lys | TET2 c.3563G>A |
| FD* | 825.06 | 165 | 8 | 102 | chr4 | 106193849 | . | G | GA | 24 | PASS | INDEL | 8010 | 866 | 25 | 0.0288684 | GA | frameshift_variant | HIGH | TET2 | c.4380dupA | p.Arg1461fs | TET2 c.4380dupA |
| FD | 986.43 | 199 | 303 | 110 | chr4 | 106164772 | . | C | T | 6 | PASS | SNP | 4671 | 747 | 8 | 0.0107095 | T | protein_protein_contact | HIGH | TET2 | c.3640C>T | p.Arg1214Trp | TET2 c.3640C>T |
| FD | 986.43 | 199 | 303 | 110 | chr4 | 106158075 | . | T | A | 30 | PASS | SNP | 2738 | 508 | 22 | 0.0433071 | A | stop_gained | HIGH | TET2 | c.3039T>A | p.Cys1013* | TET2 c.3039T>A |
| FD | 1016.42 | 172 | 12 | 176 | chr4 | 106196229 | . | T | TCATGCAGCAGTCCCAGC | 7 | PASS | INDEL | 6604 | 792 | 9 | 0.0113636 | TCATGCAGCAGTCCCAGC | frameshift_variant | HIGH | TET2 | c.4626_4642dupCATGCAGCAGTCCCAGC | p.Gln1548fs | TET2 c.4626_4642dupCATGCAGCAGTCCCAGC |
| FD | 1019.4 | 172 | 94 | 161 | chr4 | 106194066 | . | C | T | 7 | PASS | SNP | 4204 | 608 | 8 | 0.0131579 | T | stop_gained | HIGH | TET2 | c.4591C>T | p.Gln1531* | TET2 c.4591C>T |
| FD | 1088.81 | 186 | 270 | 100 | chr4 | 106157474 | . | C | G | 8 | PASS | SNP | 8129 | 1018 | 9 | 0.0088409 | G | stop_gained | HIGH | TET2 | c.2438C>G | p.Ser813* | TET2 c.2438C>G |
| CD | 1117 | 164 | 73 | 3 | chr4 | 106157069 | . | C | CA | 7 | PASS | INDEL | 5497 | 793 | 9 | 0.0113493 | CA | frameshift_variant | HIGH | TET2 | c.2034dupA | p.His679fs | TET2 c.2034dupA |
| CD | 1146.5 | 165 | 132 | 5 | chr4 | 106196374 | . | C | A | 16 | PASS | SNP | 4431 | 724 | 18 | 0.0248619 | A | stop_gained | HIGH | TET2 | c.4770C>A | p.Tyr1590* | TET2 c.4770C>A |
| FD | 1181.78 | 154 | 116 | 102 | chr4 | 106180790 | COSM87135 | G | C | 7 | PASS | SNP | 5085 | 640 | 9 | 0.0140625 | C | missense_variant | MODERATE | TET2 | c.3881G>C | p.Cys1294Ser | TET2 c.3881G>C |
| CD | 1207.44 | 172 | 193 | 6 | chr4 | 106164061 | . | C | T | 19 | PASS | SNP | 4832 | 638 | 19 | 0.0297806 | T | stop_gained | HIGH | TET2 | c.3634C>T | p.Gln1212* | TET2 c.3634C>T |
| FD | 1266.67 | 154 | 369 | 136 | chr4 | 106180835 | . | G | A | 59 | PASS | SNP | 4293 | 622 | 46 | 0.073955 | A | missense_variant | MODERATE | TET2 | c.3926G>A | p.Gly1309Asp | TET2 c.3926G>A |
| FD | 1305 | 172 | 347 | 100 | chr4 | 106196551 | . | T | G | 6 | PASS | SNP | 7347 | 1152 | 10 | 0.0086806 | G | stop_gained | HIGH | TET2 | c.4947T>G | p.Tyr1649* | TET2 c.4947T>G |
| FD | 1305 | 172 | 347 | 100 | chr4 | 106196664 | . | CT | C | 6 | PASS | INDEL | 5298 | 819 | 8 | 0.009768 | C | frameshift_variant | HIGH | TET2 | c.5061delT | p.Leu1688fs | TET2 c.5061delT |
| FD | 1421.81 | 172 | 291 | 116 | chr4 | 106156825 | . | G | T | 15 | PASS | SNP | 7077 | 894 | 19 | 0.0212528 | T | stop_gained | HIGH | TET2 | c.1789G>T | p.Glu597* | TET2 c.1789G>T |
| FD | 2061.18 | 155 | 244 | 121 | chr4 | 106157212 | . | C | T | 28 | PASS | SNP | 7583 | 1838 | 39 | 0.0212187 | T | stop_gained | HIGH | TET2 | c.2176C>T | p.Gln726* | TET2 c.2176C>T |
| FD | 2224.76 | 154 | 402 | 100 | chr4 | 106158419 | . | C | CA | 15 | PASS | INDEL | 9707 | 1444 | 21 | 0.0145429 | CA | frameshift_variant | HIGH | TET2 | c.3384dupA | p.Pro1129fs | TET2 c.3384dupA |
| CD | 2331.81 | 205 | 415 | 4 | chr4 | 106164936 | . | G | A | 23 | PASS | SNP | 3346 | 1025 | 27 | 0.0263415 | A | splice_donor_variant&intron_variant | HIGH | TET2 | c.3866+1G>A | NA | TET2 c.3866+1G>A |
| CD | 2685.15 | 166 | 372 | 6 | chr4 | 106156744 | . | ACACGAGATCTTGTG | A | 7 | PASS | INDEL | 7350 | 2712 | 17 | 0.0062684 | A | frameshift_variant | HIGH | TET2 | c.1710_1723delACGAGATCTTGTGC | p.Arg571fs | TET2 c.1710_1723delACGAGATCTTGTGC |
| * extended FD | | | | | | | | | | | | | | | | | | | | | | | |

**Supplemental Table 5. DNMT3A fitness scores and site-specific mutation rates**

| Group | DonorID | AminoAcidExchange | FitnessScore (% growth per year) | SiteSpecificMutationRate (µ x10*-9 per year) |
|---|---|---|---|---|
| FD | 253 | p.Arg882Cys | 12,30 | 1,42E-03 |
| FD | 122 | p.Arg882Cys | 12,30 | 1,42E-03 |
| FD | 42 | p.Glu733* | 12,38 | 5,40E-05 |
| FD | 244 | p.Arg882His | 13,07 | 1,88E-03 |
| FD | 31 | p.Asn838Asp | 14,59 | 4,96E-05 |
| FD | 57 | p.Phe755Ser | 14,96 | 4,96E-05 |
| FD | 291 | p.Arg366His | 7,30 | 1,88E-03 |
| FD | 321 | p.Ser770Leu | 8,04 | 1,20E-03 |
| FD | 3 | p.Arg635Trp | 8,98 | 1,42E-03 |
| FD | 136 | p.Arg635Trp | 8,98 | 1,42E-03 |
| FD | 310 | p.Arg749Cys | 9,22 | 1,20E-03 |
| FD | 3 | p.Arg771* | 9,27 | 1,88E-03 |
| FD | 273 | p.Arg771* | 9,27 | 1,88E-03 |
| FD | 18 | p.Arg736Cys | 9,42 | 1,42E-03 |
| FD | 3 | p.Trp305* | 9,88 | 5,14E-04 |
| CD | 430 | p.Met761Val | 11,66 | 1,49E-04 |
| CD | 371 | p.Arg882Cys | 12,30 | 1,42E-03 |
| CD | 72 | p.Ile705Thr | 12,32 | 1,35E-04 |
| CD | 146 | p.Ile780Thr | 12,91 | 1,35E-04 |
| CD | 132 | p.Arg882His | 13,07 | 1,88E-03 |
| CD | 183 | p.Trp860Arg | 15,45 | 1,99E-03 |
| CD | 434 | p.Trp860Arg | 15,45 | 1,99E-03 |
| CD | 127 | p.Tyr735Cys | 19,93 | 8,81E-05 |

**Supplemental Table 6. Longitudinal analysis**

| Group | UMIDepth | DonorID | DateOfDonation | NumberOfDonations | Donation | DaysBetweenDonations | CHROM | VMF | Gene_Name | HGVS.c | VMF(based on actual read counts) |
|---|---|---|---|---|---|---|---|---|---|---|---|
| FD | 1048.53 | 3 | 12.01.21 00:00 | 121 | 2 | 399 | chr2 | 0.0234043 | DNMT3A | c.2311C>T | |
| FD | 1048.53 | 3 | 12.01.21 00:00 | 121 | 2 | 399 | chr2 | 0.0080808 | DNMT3A | c.1903C>T | |
| FD | 1048.53 | 3 | 12.01.21 00:00 | 121 | 2 | 399 | chr2 | 0.0328228 | DNMT3A | c.915G>A | |
| FD | 1248.82 | 3 | 10.12.19 00:00 | 115 | 1 | 0 | chr2 | 0.0275362 | DNMT3A | c.2311C>T | |
| FD | 1248.82 | 3 | 10.12.19 00:00 | 115 | 1 | 0 | chr2 | 0.0065923 | DNMT3A | c.1903C>T | |
| FD | 1248.82 | 3 | 10.12.19 00:00 | 115 | 1 | 0 | chr2 | 0.0321285 | DNMT3A | c.915G>A | |
| FD* | 616.36 | 8 | 05.05.21 00:00 | 106 | 2 | 512 | chr2 | 0.0595483 | DNMT3A | c.914G>A | |
| FD* | 616.36 | 8 | 05.05.21 00:00 | 106 | 2 | 512 | chr4 | 0.0495726 | TET2 | c.4380dupA | |
| FD* | 825.06 | 8 | 10.12.19 00:00 | 102 | 1 | 0 | chr2 | 0.0210526 | DNMT3A | c.914G>A | |
| FD* | 825.06 | 8 | 10.12.19 00:00 | 102 | 1 | 0 | chr4 | 0.0288684 | TET2 | c.4380dupA | |
| FD | 805.95 | 11 | 10.12.19 00:00 | 112 | 1 | 0 | chr2 | 0.0276596 | DNMT3A | c.2323-1G>T | |
| FD | 2056.83 | 11 | 20.04.21 00:00 | 115 | 2 | 497 | chr2 | 0.0448679 | DNMT3A | c.2323-1G>T | |
| FD | 699.5 | 31 | 11.12.19 00:00 | 138 | 1 | 0 | chr2 | 0.0151515 | DNMT3A | c.2512A>G | |
| FD | 699.5 | 31 | 11.12.19 00:00 | 138 | 1 | 0 | chr2 | 0.0751174 | DNMT3A | c.2193_2195delCTT | |
| FD | 3571.48 | 31 | 09.12.20 00:00 | 143 | 2 | 364 | chr2 | 0.0310289 | DNMT3A | c.2512A>G | |
| FD | 3571.48 | 31 | 09.12.20 00:00 | 143 | 2 | 364 | chr2 | 0.0788127 | DNMT3A | c.2193_2195delCTT | |
| FD | 774.9 | 42 | 11.12.19 00:00 | 124 | 1 | 0 | chr2 | 0.0140351 | DNMT3A | c.1040T>C | |
| FD | 774.9 | 42 | 11.12.19 00:00 | 124 | 1 | 0 | chr2 | 0.091858 | DNMT3A | c.2197G>T | |
| FD | 1315.09 | 42 | 10.02.21 00:00 | 128 | 2 | 427 | chr2 | 0.0145985 | DNMT3A | c.1040T>C | |
| FD | 1315.09 | 42 | 10.02.21 00:00 | 128 | 2 | 427 | chr2 | 0.11323 | DNMT3A | c.2197G>T | |
| FD | 802.71 | 45 | 11.12.19 00:00 | 101 | 1 | 0 | chr4 | 0.0127389 | TET2 | c.2480delT | |
| FD | 1079.14 | 45 | 10.02.21 00:00 | 105 | 2 | 427 | chr4 | 0.0108803 | TET2 | c.2480delT | |
| FD | 1056.89 | 57 | 11.12.19 00:00 | 122 | 1 | 0 | chr2 | 0.0093857 | DNMT3A | c.2264T>C | |
| FD | 1130.58 | 57 | 08.12.20 00:00 | 126 | 2 | 363 | chr2 | 0.0099458 | DNMT3A | c.2264T>C | |
| FD | 1019.4 | 94 | 15.01.20 00:00 | 161 | 1 | 0 | chr4 | 0.0131579 | TET2 | c.4591C>T | |
| FD | 1411.76 | 94 | 24.02.21 00:00 | 165 | 2 | 406 | chr4 | 0.0214944 | TET2 | c.4591C>T | |
| FD | 1308.2 | 136 | 18.05.20 00:00 | 111 | 1 | 0 | chr2 | 0.0082902 | DNMT3A | c.1903C>T | |
| FD | 1481.27 | 136 | 17.05.21 00:00 | 116 | 2 | 364 | chr2 | 0.0067797 | DNMT3A | c.1903C>T | |
| FD | 1116.78 | 200 | 08.03.21 00:00 | 115 | 2 | 287 | chr2 | 0.0200445 | DNMT3A | c.1667+1G>A | |
| FD | 1432.01 | 200 | 25.05.20 00:00 | 111 | 1 | 0 | chr2 | 0.0215664 | DNMT3A | c.1667+1G>A | |
| FD | 919.07 | 248 | 27.05.20 00:00 | 111 | 1 | 0 | chr2 | 0.0286807 | DNMT3A | c.1434delG | |
| FD | 1094.42 | 248 | 19.01.21 00:00 | 115 | 2 | 237 | chr2 | 0.0444785 | DNMT3A | c.1434delG | |
| FD | 1057.75 | 310 | 06.01.21 00:00 | 103 | 2 | 218 | chr2 | 0.0074129 | DNMT3A | c.2245C>T | |
| FD | 1064.53 | 310 | 02.06.20 00:00 | 100 | 1 | 0 | chr2 | 0.0069009 | DNMT3A | c.2245C>T | |
| FD | 1125.23 | 321 | 02.06.20 00:00 | 143 | 1 | 0 | chr2 | 0.0773196 | DNMT3A | c.2309C>T | |
| FD | 1356.21 | 321 | 04.06.21 00:00 | 149 | 2 | 367 | chr2 | 0.0569444 | DNMT3A | c.2309C>T | |
| FD | 1108.1 | 352 | 12.04.21 00:00 | 151 | 2 | 308 | chr2 | 0.0514019 | DNMT3A | c.2201T>C | |
| FD | 1387.65 | 352 | 08.06.20 00:00 | 147 | 1 | 0 | chr2 | 0.0440613 | DNMT3A | c.2201T>C | |
| FD | 802.55 | 369 | 02.03.21 00:00 | 140 | 2 | 267 | chr4 | 0.1003788 | TET2 | c.3863G>A | |
| FD | 1266.67 | 369 | 08.06.20 00:00 | 136 | 1 | 0 | chr4 | 0.073955 | TET2 | c.3863G>A | |
| FD | 2224.76 | 402 | 05.06.20 00:00 | 100 | 1 | 0 | chr4 | 0.0145429 | TET2 | c.3384dupA | |
| FD | 1775.54 | 402 | 04.06.21 00:00 | 105 | 2 | 364 | chr4 | 0.0099010 | TET2 | c.3384dupA | |
| CD | 1867.19 | 69 | 08.01.20 00:00 | 10 | 1 | 0 | chr2 | 0.0077419 | DNMT3A | c.2194_2201delTTTGAGTT | |
| CD | 1677.33 | 69 | 16.06.21 00:00 | 14 | 2 | 520 | chr2 | 0.0106007 | DNMT3A | c.2194_2201delTTTGAGTT | |
| CD | 1029.25 | 75 | 07.01.20 00:00 | 7 | 1 | 0 | chr2 | 0.0192593 | DNMT3A | c.1051dupT | |
| CD | 1323.62 | 75 | 04.03.21 00:00 | 11 | 2 | 422 | chr2 | 0.021645 | DNMT3A | c.1051dupT | |
| CD | 896.57 | 177 | 25.05.20 00:00 | 3 | 1 | 0 | chr2 | 0.0155521 | DNMT3A | c.2521A>G | |
| CD | 896.57 | 177 | 25.05.20 00:00 | 3 | 1 | 0 | chr2 | 0.0636833 | DNMT3A | c.2132delA | |
| CD | 1208.98 | 177 | 30.06.21 00:00 | 7 | 2 | 401 | chr2 | 0.0168675 | DNMT3A | c.2521A>G | |
| CD | 1208.98 | 177 | 30.06.21 00:00 | 7 | 2 | 401 | chr2 | 0.1228346 | DNMT3A | c.2132delA | |
| CD | 850.12 | 371 | 11.03.21 00:00 | 5 | 2 | 275 | chr2 | 0.0204878 | DNMT3A | c.2644C>T | |
| CD | 1071.62 | 371 | 09.06.20 00:00 | 3 | 1 | 0 | chr2 | 0.0131805 | DNMT3A | c.2644C>T | |
| CD | 1629.93 | 415 | 10.02.21 00:00 | 5 | 2 | 245 | chr4 | 0.0291153 | TET2 | c.3866+1G>A | |
| CD | 2331.81 | 415 | 10.06.20 00:00 | 4 | 1 | 0 | chr4 | 0.0263415 | TET2 | c.3866+1G>A | |
| CD | 1223.71 | 444 | 26.06.20 00:00 | 2 | 1 | 0 | chr2 | 0.0077419 | DNMT3A | c.2255_2257delTCT | 0.006 |
| CD | 1125.80 | 444 | 10.09.21 00:00 | 7 | 2 | 441 | chr2 | 0 | DNMT3A | c.2255_2257delTCT | 0.002 |
| * extended FD | | | | | | | | | | | |

**Supplemental Table 7. Differential blood count analysis of Buffy Coats**

| Bzffy Coat ID | | 7,032E+10 | 7,032E+10 | 7,042E+10 | 7,042E+10 | 7,012E+10 | 7,042E+10 | 7,042E+10 | 7,032E+10 | 7,042E+10 | 7,042E+10 | 7,032E+10 | 7,032E+10 | 7,032E+10 | 7,032E+10 | 7,042E+10 | 7,042E+10 | 7,032E+10 | 7,032E+10 | 7,042E+10 | 7,032E+10 | 7,042E+10 |
|---|---|---|---|---|---|---|---|---|---|---|---|---|---|---|---|---|---|---|---|---|---|---|
| | Hb(g/dl) | 17,0 | 17,1 | 17,2 | 19,2 | 20,3 | 18,8 | 18,2 | 20,1 | 18,1 | 19,1 | 20,5 | 16,8 | 19,0 | 21,3 | 18,2 | 15,9 | 19,2 | 18,0 | 16,9 | 17,7 | 18,7 |
| | Hct(%) | 56,6 | 58,5 | 58,1 | 62,6 | 65,3 | 60,0 | 56,3 | 67,1 | 62,5 | 65,3 | 67,1 | 56,4 | 64,1 | 68,5 | 62,3 | 54,3 | 64,5 | 60,7 | 56,2 | 61,2 | 65,2 |
| | MCV(fl) | 103,5 | 100,2 | 106,8 | 104,2 | 108,3 | 97,7 | 99,8 | 104,0 | 104,0 | 105,7 | 98,8 | 105,0 | 106,3 | 103,9 | 100,0 | 100,9 | 107,5 | 103,9 | 109,6 | 103,7 | 97,3 |
| | MCH(pg) | 31,1 | 29,3 | 31,6 | 31,9 | 33,7 | 30,6 | 32,3 | 31,2 | 30,1 | 30,9 | 30,2 | 31,3 | 31,5 | 32,3 | 29,2 | 29,6 | 32,0 | 30,8 | 32,9 | 30,0 | 27,9 |
| | MCHC(g/dl) | 30,0 | 29,2 | 29,6 | 30,7 | 31,1 | 31,3 | 32,3 | 30,0 | 29,0 | 29,2 | 30,6 | 29,8 | 29,6 | 31,1 | 29,2 | 29,3 | 29,8 | 29,7 | 30,1 | 28,9 | 28,7 |
| | RDW-CV(%) | 13,9 | 18,7 | 13,3 | 14,5 | 14,3 | 14,8 | 14,4 | 14,2 | 14,2 | 14,5 | 14,4 | 13,1 | 14,1 | 15,4 | 14,1 | 12,9 | 13,7 | 14,1 | 13,4 | 13,4 | 16,5 |
| | Plt(x10$^3$/µl) | 3049 | 2966 | 2491 | 3095 | 2643 | 2969 | 800 | 2599 | 2643 | 3099 | 3428 | 2989 | 3209 | 3724 | 3422 | 3855 | 3522 | 1993 | 1930 | 3614 | 3849 |
| Day 1 | MPV(fl) | 10,0 | 9,3 | 8,8 | 10,7 | 9,7 | 9,8 | 8,8 | 10,1 | 10,3 | 9,2 | 8,5 | 9,4 | 10,2 | 8,8 | 9,6 | 9,0 | 8,9 | 10,3 | 11,4 | 9,7 | 8,4 |
| | PDW(fl) | 13,2 | 11,6 | 10,4 | 14,3 | 12,4 | 12,9 | 10,2 | 13,5 | 13,5 | 11,4 | 10,5 | 11,8 | 13,4 | 11,2 | 12,3 | 10,9 | 10,8 | 13,5 | 15,8 | 12,6 | 10,3 |
| | WBC (x10$^3$/µ | 65,58 | 61,57 | 54,05 | 53,02 | 61,59 | 87,05 | 19,23 | 56,40 | 48,54 | 58,45 | 63,47 | 44,10 | 77,12 | 81,24 | 79,49 | 62,22 | 60,33 | 82,09 | 70,90 | 44,47 | 89,20 |
| | Neut(%) | 31,45 | 22,85 | 24,53 | 21,62 | 41,26 | 52,03 | 6,87 | 23,04 | 22,47 | 19,54 | 28,06 | 12,99 | 31,91 | 44,48 | 29,83 | 23,30 | 25,84 | 39,92 | 34,42 | 7,01 | 33,64 |
| | Lymph(%) | 27,01 | 32,12 | 20,38 | 24,21 | 15,28 | 26,63 | 9,42 | 25,48 | 18,96 | 27,29 | 24,88 | 24,95 | 36,44 | 29,11 | 34,89 | 28,74 | 29,36 | 33,44 | 25,44 | 31,62 | 45,00 |
| | Mono(%) | 4,92 | 5,05 | 7,27 | 5,83 | 3,61 | 6,97 | 1,89 | 6,65 | 6,15 | 10,96 | 9,54 | 5,68 | 7,43 | 5,53 | 12,36 | 8,77 | 4,41 | 7,25 | 7,06 | 4,99 | 9,19 |
| | Eo(%) | 1,67 | 0,98 | 1,28 | 0,41 | 1,03 | 0,40 | 0,90 | 0,11 | 0,35 | 0,23 | 0,59 | 0,11 | 0,44 | 1,40 | 1,22 | 0,96 | 0,30 | 0,57 | 0,45 | 0,61 | 0,77 |
| | Baso(%) | 0,53 | 0,57 | 0,59 | 0,95 | 0,41 | 1,02 | 0,15 | 1,12 | 0,61 | 0,43 | 0,40 | 0,37 | 0,90 | 0,72 | 1,19 | 0,45 | 0,42 | 0,91 | 0,53 | 0,24 | 0,60 |
| | IG(%) | 0,24 | 0,20 | 0,20 | 0,14 | 0,30 | 0,72 | 0,02 | 0,19 | 0,14 | 0,17 | 0,24 | 0,12 | 0,37 | 0,48 | 0,26 | 0,20 | 0,21 | 0,90 | 0,50 | 0,06 | 0,64 |
| | Hb(g/dl) | 16,6 | 17,0 | 17,3 | 18,7 | 19,3 | 18,7 | 18,0 | 19,7 | 17,9 | 19,1 | 20,6 | 16,9 | 19,0 | 21,2 | 18,1 | 15,9 | 19,5 | 17,8 | 16,9 | 17,9 | 18,9 |
| | Hct(%) | 59,3 | 62,0 | 61,2 | 67,2 | 66,1 | 65,1 | 57,4 | 68,7 | 65,4 | 68,2 | 71,1 | 61,5 | 69,8 | 73,4 | 67,5 | 60,8 | 68,4 | 63,5 | 60,3 | 66,7 | 70,4 |
| | MCV(fl) | 112,1 | 108,6 | 112,9 | 105,7 | 115,8 | 106,2 | 102,7 | 112,1 | 111,0 | 112,2 | 105,5 | 108,5 | 108,4 | 109,1 | 103,7 | 107,0 | 115,5 | 112,0 | 111,3 | 107,2 | 106,8 |
| | MCH(pg) | 31,4 | 29,8 | 31,9 | 29,4 | 33,8 | 30,5 | 32,2 | 32,1 | 30,4 | 31,4 | 30,6 | 29,8 | 29,5 | 31,5 | 27,8 | 28,0 | 32,9 | 31,4 | 31,2 | 28,8 | 28,7 |
| | MCHC(g/dl) | 28,0 | 27,4 | 28,3 | 27,8 | 29,2 | 28,7 | 31,4 | 28,7 | 27,4 | 28,0 | 29,0 | 27,5 | 27,2 | 28,9 | 26,8 | 26,2 | 28,5 | 28,0 | 28,0 | 26,8 | 26,8 |
| | RDW-CV(%) | 13,7 | 18,9 | 13,4 | 20,0 | 13,3 | 15,0 | 14,8 | 14,4 | 14,1 | 14,2 | 13,7 | 17,5 | 17,8 | 16,9 | 17,7 | 20,8 | 13,9 | 14,0 | 17,8 | 18,4 | 16,7 |
| | | | | | | | | | 4,00 | | | | | | | | | | | | | |
| | Plt(x10$^3$/µl) | 3417 | 4275 | 2860 | 33 | 3026 | 3372 | 902 | 2773 | 2750 | 3494 | 3966 | 3428 | 3525 | 4303 | 3920 | 4484 | 4153 | 2120 | 2033 | 4043 | 4598 |
| Day 2 | MPV(fl) | 11,5 | 10,9 | 9,8 | 12,4 | 10,5 | 11,5 | 8,9 | 11,6 | 11,3 | 10,5 | 9,6 | 11,4 | 11,8 | 10,4 | 11,6 | 11,7 | 10,3 | 12,1 | 12,7 | 11,6 | 10,4 |
| | PDW(fl) | 16,7 | 15,4 | 12,6 | 18,4 | 14,3 | 16,6 | 10,3 | 17,2 | 16,5 | 14,4 | 12,6 | 16,1 | 17,4 | 14,6 | 17,0 | 16,8 | 13,5 | 18,2 | 19,9 | 16,9 | 14,2 |
| | WBC (x10$^3$/µ | 65,37 | 60,94 | 51,00 | 53,15 | 61,22 | 88,96 | 18,49 | 56,22 | 45,54 | 58,45 | 64,34 | 44,12 | 75,77 | 77,61 | 78,72 | 61,66 | 60,31 | 83,72 | 69,18 | 45,67 | 88,73 |
| | Neut(%) | 31,60 | 22,11 | 22,78 | 21,91 | 40,21 | 51,24 | 6,66 | 22,46 | 20,18 | 19,88 | 28,69 | 12,55 | 30,77 | 41,27 | 29,19 | 23,18 | 24,83 | 39,77 | 35,90 | 7,07 | 32,80 |
| | Lymph(%) | 25,54 | 30,83 | 18,01 | 23,00 | 15,42 | 27,53 | 9,02 | 23,49 | 17,09 | 25,16 | 24,32 | 24,35 | 36,02 | 26,39 | 33,48 | 26,13 | 29,83 | 33,80 | 25,14 | 31,41 | 42,92 |
| | Mono(%) | 5,51 | 5,75 | 7,44 | 6,69 | 3,68 | 7,52 | 1,80 | 8,45 | 6,92 | 12,60 | 10,40 | 6,49 | 7,52 | 6,97 | 12,45 | 10,65 | 4,30 | 7,66 | 6,80 | 6,10 | 10,72 |
| | Eo(%) | 1,50 | 0,98 | 1,16 | 0,33 | 1,00 | 0,44 | 0,83 | 0,14 | 0,31 | 0,28 | 0,42 | 0,09 | 0,37 | 1,10 | 1,20 | 0,80 | 0,24 | 0,62 | 0,43 | 0,70 | 0,75 |
| | Baso(%) | 1,22 | 1,27 | 1,61 | 1,22 | 0,91 | 2,23 | 0,18 | 1,68 | 1,04 | 0,53 | 0,87 | 0,64 | 1,08 | 1,88 | 2,40 | 0,69 | 1,11 | 1,87 | 0,91 | 0,39 | 1,54 |
| | IG(%) | 0,44 | 0,68 | 0,58 | 0,52 | 0,65 | 0,81 | 0,05 | 0,61 | 0,35 | 0,78 | 0,54 | 0,35 | 0,57 | 0,61 | 0,66 | 0,61 | 0,27 | 1,14 | 0,62 | 0,12 | 1,02 |
| | Hb(g/dl) | 18,2 | 17,2 | 17,0 | 18,7 | 19,2 | 18,4 | 16,3 | 18,7 | 18,0 | 18,9 | 20,0 | 16,8 | 19,0 | 21,1 | 17,3 | 15,8 | 19,3 | 17,8 | 16,9 | 17,7 | 18,6 |
| | Hct(%) | 67,1 | 62,3 | 60,3 | 68,3 | 66,8 | 64,4 | 52,8 | 67,4 | 66,0 | 68,4 | 70,5 | 60,4 | 69,4 | 74,1 | 63,5 | 59,5 | 69,0 | 65,3 | 61,4 | 66,4 | 70,4 |
| | MCV(fl) | 107,5 | 109,1 | 114,2 | 105,9 | 112,6 | 107,5 | 104,6 | 107,5 | 107,8 | 112,7 | 106,5 | 112,9 | 107,9 | 109,6 | 108,9 | 112,9 | 111,7 | 107,8 | 112,5 | 106,4 | 107,3 |
| | MCH(pg) | 29,2 | 30,1 | 32,2 | 29,0 | 32,4 | 30,7 | 32,3 | 29,8 | 29,4 | 31,1 | 30,2 | 31,4 | 29,5 | 31,2 | 29,7 | 30,0 | 31,2 | 29,4 | 31,0 | 28,4 | 28,4 |
| | MCHC(g/dl) | 27,1 | 27,6 | 28,2 | 27,4 | 28,7 | 28,6 | 30,9 | 27,7 | 27,3 | 27,6 | 28,4 | 27,8 | 27,4 | 28,5 | 27,2 | 26,6 | 28,0 | 27,3 | 27,5 | 26,7 | 26,4 |
| | RDW-CV(%) | 19,4 | 18,6 | 13,8 | 20,9 | 16,8 | 15,7 | 12,0 | 17,6 | 16,9 | 14,8 | 14,6 | 13,2 | 18,8 | 17,8 | 13,7 | 13,2 | 16,5 | 17,2 | 19,3 | 19,4 | 16,7 |
| | Plt(x10$^3$/µl) | 3320 | 3818 | 2959 | 3115 | 3079 | 3022 | 898 | 2706 | 2812 | 3288 | 3855 | 3279 | 3204 | 4240 | 3588 | 3878 | 4141 | 2020 | 1991 | 3801 | 3916 |
| Day 3 | MPV(fl) | 11,9 | 11,1 | 10,2 | 12,9 | 11,1 | 12,0 | 9,9 | 12,0 | 11,6 | 11,2 | 9,9 | 11,5 | 12,3 | 10,7 | 11,5 | 11,7 | 10,5 | 12,5 | 13,2 | 12,0 | 11,1 |
| | PDW(fl) | 17,9 | 15,7 | 13,3 | 19,6 | 15,5 | 18,2 | 12,0 | 17,9 | 16,8 | 16,1 | 13,0 | 16,3 | 18,7 | 14,9 | 16,9 | 16,8 | 14,4 | 19,4 | 21,0 | 17,9 | 15,9 |
| | WBC (x10$^3$/µ | 68,72 | 61,05 | 51,11 | 53,56 | 62,35 | 86,51 | 17,75 | 53,99 | 46,15 | 56,96 | 61,87 | 43,79 | 74,26 | 76,96 | 75,08 | 61,80 | 58,52 | 81,66 | 68,46 | 44,90 | 89,50 |
| | Neut(%) | 32,71 | 20,52 | 25,02 | 22,02 | 41,42 | 50,36 | 6,65 | 20,87 | 21,58 | 20,22 | 29,49 | 13,44 | 32,04 | 41,33 | 29,93 | 25,29 | 24,10 | 39,66 | 34,39 | 8,16 | 33,10 |
| | Lymph(%) | 25,82 | 30,17 | 16,55 | 22,03 | 14,59 | 25,11 | 8,87 | 20,67 | 16,75 | 23,04 | 21,34 | 23,57 | 33,88 | 23,38 | 29,85 | 25,87 | 27,00 | 31,03 | 22,76 | 29,46 | 41,08 |
| | Mono(%) | 5,47 | 6,62 | 5,67 | 6,96 | 3,12 | 7,09 | 1,19 | 9,48 | 5,91 | 12,35 | 8,63 | 5,39 | 6,19 | 8,06 | 10,69 | 8,82 | 4,99 | 7,43 | 7,62 | 5,89 | 11,91 |
| | Eo(%) | 1,54 | 0,86 | 1,12 | 0,36 | 0,87 | 0,42 | 0,75 | 0,12 | 0,30 | 0,22 | 0,44 | 0,09 | 0,37 | 1,01 | 1,07 | 0,76 | 0,22 | 0,57 | 0,38 | 0,61 | 0,75 |
| | Baso(%) | 3,18 | 2,88 | 2,75 | 2,19 | 2,35 | 3,53 | 0,29 | 2,85 | 1,61 | 1,13 | 1,97 | 1,30 | 1,78 | 3,18 | 3,54 | 1,06 | 2,21 | 2,97 | 3,31 | 0,78 | 2,66 |
| | IG(%) | 1,25 | 0,59 | 2,08 | 1,09 | 1,66 | 11,70 | 0,43 | 0,92 | 1,09 | 1,70 | 1,48 | 1,17 | 1,60 | 1,18 | 1,70 | 1,53 | 0,53 | 1,63 | 1,31 | 0,60 | 1,50 |

**Supplemental Table 8. Digital droplet PCR Assays**

| AssayName | AssayID | AminoAcidExchange | NucleotideMutation | COSMICID |
|---|---|---|---|---|
| DNMT3A p.W305* c.914G>A | dHsaMDS311847198 | p.W305* | c.914G>A | COSM1169636 |
| DNMT3A p.F543delF / DNMT3A p.F731del c.2191_2193delTTC | dHsaMDS549125373 / dHsaMDS802387875 | p.F732del (old: p.F543delF) | c.2191_2193delTTC | COSM99742 |
| DNMT3A p.E733* c.2197C>T | dHsaMDS529169084 | pE733* | c.2197C>T | |
| DNMT3A p.L347P c.1040T>C | dHsaMDS126094721 | p.L347P | c.1040T>C | COSM5944978 |
| DNMT3A p.R882H c.2645G>A | dHsaMDV2010089 | p.R882H | c.2645G>A | COSM52944 |
| DNMT3A p.R882C c.2644C>T | dHsaMDS475153762 | p.R882C | c.2644C>T | COSM53042 |

**Supplemental Table 9. CRISPR guides and HDR donor templates**

| CHROM | HGVS.c | HGVS.p | sgRNA | Donor template | MiSeq primers |
|---|---|---|---|---|---|
| chr2 | c.2197G>T | p.Glu733* | TCCGACCTCTCAGAGGGCAC | GCGATCATCTCCCTCCTTGGGCCGCGCATCATGCAGGAGGCGGTAGAACT<br>AAAAGAAGAGCCGGCCAGTGCCCTCTGAGAGGTCGGAAGAGAAAGCCATC | **FWD:** TCGTCGGCAGCGTCAGATGTGTATAAGAGACAGGTTGCTGGCTATACCTCGAG.<br>**REV:** GTCTCGTGGGCTCGGAGATGTGTATAAGAGACAGGGATATTTCTGCCCTGGGAC. |
| chr2 | c.914G>A | p.Trp305* | GGAAACTGCGGGGCTTCTCC | AGCTGCTCGGCTCCGGCCCGTCATCCACCAAGACACAATGCGGCCTGGCC<br>ACTAGGAGAAGCCCCGCAGTTTCCCCCACACCAGCTCCCCAATGCCAAAG | **FWD:** TCGTCGGCAGCGTCAGATGTGTATAAGAGACAGCTACTGCCAAACCCCACAAC.<br>**REV:** GTCTCGTGGGCTCGGAGATGTGTATAAGAGACAGCTCGTGACCACTGTGTAATG. |
| chr2 | c.1988delC | p.Ser663fs | GGACCGCTACATTGCCTCGG | TTCCCCTGGTGCCGCACCATGCCCACCGTGATGGAGTCCTCACACACCTC<br>CAGGCAATGTAGCGGTCCACCTGAATGCCCAAGTCCTTCAGCACCAGGA | **FWD:** TCGTCGGCAGCGTCAGATGTGTATAAGAGACAGCACAGCATGGACATACATGC.<br>**REV:** GTCTCGTGGGCTCGGAGATGTGTATAAGAGACAGATGGGTCCTGTTGGCCAGCA. |
| chr2 | c.2644C>T (R882C) | p.Arg882Cys | CCTCGCCAAGCGGCTCATGT | GATGACTGGCACGCTCCATGACCGGCCCAGCAGTCTCTGTCTCGCTAAGCAGC<br>TCATGTTCGAGACGTCAGTATAGTGGACTGGGAAACCAAATACCCTG | **FWD:** TCGTCGGCAGCGTCAGATGTGTATAAGAGACAGAGGAGTTGGTGGGTGTGAGT.<br>**REV:** GTCTCGTGGGCTCGGAGATGTGTATAAGAGACAGCACGCAAAATACTCCTTCAGC. |
| chr2 | c.2645G>A (R882H) | p.Arg882His | CCTCGCCAAGCGGCTCATGT | GATGACTGGCACGCTCCATGACCGGCCCAGCAGTCTCTGTCTCGCTAAGTGGC<br>TCATGTTCGAGACGTCAGTATAGTGGACTGGGAAACCAAATACCCTG | **FWD:** TCGTCGGCAGCGTCAGATGTGTATAAGAGACAGAGGAGTTGGTGGGTGTGAGT.<br>**REV:** GTCTCGTGGGCTCGGAGATGTGTATAAGAGACAGCACGCAAAATACTCCTTCAGC. |

### Supplemental Methods

#### Healthy Blood Donors

Buffy coats, waste products of component preparation from whole blood donations, of selected healthy male volunteer blood donors donating between December 2019 and June 2020 as well as December 2020 and November 2021 (for consecutive samples) at the German Red Cross Blood Service Baden-Württemberg-Hessen were used for the study. All donors signed an informed consent allowing for anonymous processing of the samples. Database query parameter were set to select male individuals over the age of 60 with greater than 100 (frequent blood donors) or fewer than 10 (infrequent control donors) whole blood donations. Buffy coats produced in the course of processing of erythrocyte concentrates meeting these criteria were flagged by the IT system during blood processing. Subsequent verification of the donor characteristics showed that 212 out of 218 samples matched the set criteria. Further restriction with regard to inclusion of the donors was related to the quality of the sequencing (coverage depth and number of variants detected per sample) as described below under "Analysis of the variants". Based on sequencing metrics 4 additional donors (3x UMI Depth < 400; 1x 225 total variants per sample) were excluded. Thus 105 donors in the frequent donor (FD) cohort and 103 donors from the control donor (CD) cohort, were included as **main cohorts** (see Table 1 and Supplemental Table 1). These main cohorts were used for all statistical comparisons of FDs and CDs. All donor IDs are research IDs introduced in course of the study for the purpose of data analysis exclusively and do not allow an identification of the donors to anyone outside the research group.

In two of the "wrongly" processed and analyzed samples, from donor #8 (age 51-55 / 102 donations) and donor #17 (age 61-65 / 80 donations) mutations in CH drivers *DNMT3A* and *TET2* were identified. Both donors were therefore kept as part of the **extended** FD cohort despite being formally too young (donor #8) or having donated too few whole blood units for inclusion (donor #17) since they were of an age where CH can be detected at the set depth of the sequencing and met a general definition of frequent donor. Variants from these extended FD cohort donors were only considered in qualitative assessment of the variants, including longitudinal and functional analysis as well as lineage tracing.

#### Samples

##### Bulk / Whole PBMC Samples

Buffy coats (BC) were kept at RT for up to 3 days prior to shipment and processing at the German Cancer Research Center in Heidelberg. Before initiation of the study, we had verified that short term storage prior to freezing the cell pellet did not affect the leukocyte composition of the samples (see supplemental Table 7) and therefore did not introduce a bias in case of a different penetrance of certain mutations in different lineages such as have been reported e.g. for *DNMT3A* and *TET2* mutations[1].

5-10 ml BC suspension were washed with PBS and the cell pellet then spun down again to remove excessive plasma prior to freezing it at -80 °C. BC cell pellets were thawed on ice immediately prior to DNA isolation.

**Selected fractions**

For donors' consecutive sample analysis, BCs were obtained within 24 hours of whole blood donation. Whole PBMC sample was generated and processed as described above. The remaining sample (30-50 ml) was washed and subjected to Ficoll density centrifugation to isolate PBMC for subsequent freezing of live cells. On the days of sorting cryopreserved cells were gently thawed, washed and stained with anti-human CD3, CD14, CD19, CD34 and CD45 antibodies. Sorting of the cell fractions of interest was performed based on following immunophenotypes: T cells: $CD45^{high}CD34^{neg}CD3^{pos}CD14^{neg}CD19^{neg}$, B cells: $CD45^{high}CD34^{neg}CD3^{neg}CD14^{neg}CD19^{pos}$, Monocytes: $CD45^{high}CD34^{neg}CD3^{neg}CD14^{pos}CD19^{neg}$ and HSPC: $CD45^{dim}CD34^{pos}CD3^{neg}CD14^{neg}CD19^{neg}$. Cells were sorted into PBS/BSA, spun down and frozen as pellets at -80 °C until immediately prior to DNA isolation.

**DNA isolation**

DNA isolated was performed as per manufacturer's instructions using Qiagen DNA isolation kits: QiaAMP DNA Blood Maxi Kit (for up to 1 ml BC cell pellet), QiaAMP DNA Mini kit (for up to 200 µl BC cell pellet and more than 200K sorted cells) and QiaAMP DNA Micro kit (for fewer than 200K sorted cells).

**Library preparation and Sequencing**

Library preparation for targeted sequencing of PBMC samples was performed using the Human Myeloid Neoplasms Panel (Qiagen) that covers 141 genes and a total of 436 kilobase pairs. Per sample, 40 ng of genomic DNA were processed according to manufacturer's instructions to obtain dual indexed, molecularly barcoded (unique molecular barcodes, UMI) libraries. Library quality and size were assessed using Agilent 2100 Bioanalyzer. Quantitative verification was performed using qPCR (QIASeq Library Quant Assay Kit, Qiagen) and Qubit dsDNA HS Assay (Life Technologies). Sequencing was performed on an Illumina NextSeq 550 sequencer, with an average UMI based coverage of 1000x. Raw sequencing data along with the metadata of the analyzed cohorts will be submitted to the European Genome-Phenome Archive (EGA), hosted by the European Bioinformatics Institute and Centre for Genomic Regulation.

**Analysis of the variants**

Sequencing reads were mapped and annotated using the Qiagen web-based tool for QiaSeq Targeted DNA Enrichment Variant Calling[2]. Read processing pipeline along with the applied variant caller have been described previously[2] and are available at https://github.com/qiaseq/qiaseq-dna under GNU Affero General Public License v3.0. An average of 170 mutations were called per sample. Full lists of variant calls will be submitted to the GEO database. Following criteria were subsequently applied to account for sequencing artefacts as well as to reduce the variant lists to CHIP relevant mutations: Samples with an average UMI depth of less than 400x and an average number of variants per sample higher than 210 were excluded from the analysis. Only variants with a VAF of $\leq 0.4$ were extracted to

exclude germline variants. VAF values determined by the QiaSeq pipeline were used for all analysis except for the longitudinal sample (donation #2) from donor #444, where the read count ratio extracted from the Integrative Genomic Viewer (IGV, https://software.broadinstitute.org/software/igv/) were used. Synonymous variants, variants predicted to have a low effect along with common SNPs were excluded. The pipeline specific quality parameters "Filter" and "RepRegion" were set to "PASS" and "NA", respectively. Furthermore, given the size of the cohort, variants (same gene and position) found to occur more than 10 times were considered to be panel artefacts and excluded from the final analysis. Variants included in the final list of mutations had a VAF between 0.005 and 0.37 % and variant allele coverage of an average of 32 UMI family-based reads. Analyses were run in R software, v 4.0.1. Overall pathogenicity scoring of mutations was performed using the Combined Annotation Dependent Depletion (CADD) tool[3,4]. **C**OSMIC database[5] was used for manual curation of the variants.
Lollipop plots for DNMT3A and TET2 were generated using the lolliplot function from the R package trackViewer. Spliceosome mutations were excluded. Protein domain annotations for the plots were downloaded from https://genome.ucsc.edu/. Heatmap plots of the epigenetic modifier mutation frequencies were generated using the R package ggplot2.

**Digital Droplet PCR (ddPCR)**
Digital droplet PCR was performed for validation of the targeted sequencing as well as when screening for presence of selected mutations in specific cell fractions of the sample. All assays were designed and purchased from Bio-Rad. ddPCR Supermix for Probes (No dUTP, Bio-Rad) was used for all reactions. Assay IDs are listed in the Supplemental Table 8. Reactions were set up according to manufacturer's instructions with 5-50 ng genomic DNA as input and annealing / extension temperatures of 53-55 °C for 40 cycles. Bio-Rad QX200 Droplet Digital PCR System was used for droplet generation and analysis of the samples.

**Flow cytometry analysis and cell sorting**
All experiments were analyzed at the Flow Cytometry core facility of The Francis Crick Institute using the LSR FORTESSA (BD Biosciences) equipped with a 488-nm laser, a 561-nm laser, a 633-nm laser, and a 405-nm laser. For sorting, cell suspensions were filtered through a 35-µm nylon mesh (Falcon, Cat# 352235) and sorted in a BD FACS FUSION cell sorter equipped with 488-nm, 561-nm, 633-nm, and 405-nm lasers. The antibodies used were: CD45-FITC (clone HI30, Biolegend), CD34- PerCP-Cy5.5 (clone 8G12, BD Pharmingen) and CD38-PECy7 (clone HIT2, BD Pharmingen) for sorting of HSPCs and CD33-PE (clone P67.6, Biolegend), CD19-APCCy7 (clone HIB19, Biolegend), CD71-APC (clone OKT9, eBioscience) and CD235a-FITC (clone HIR2, BD Pharmingen) for flow cytometry analysis. Exclusion of dead cells was done by staining with the fluorescent dye DAPI (1 µg/ml; BD Biosciences, Cat# 564907) and gating out the positive cells. All experiments were analyzed with FACSDiva 6.2 (BD Biosciences) and FCS Express 7 software.

**Sample preparation for DNA sequencing**

Sorted cells were pelleted and DNA was extracted using EZNA Tissue DNA kit (Omega Bio-tek). Targeted sequencing to the region of interest was performed after PCR amplification using the corresponding primers listed in the supplemental Table 10.

***In silico* structural analysis**

Models were generated using homology modelling on SWISS-MODEL[6,7] based on the crystal structure of DNMT3A available on PDB under the alias 5YX2[8]. UCSF ChimeraX[9] and PyMOL[10] were used for model visualization.

**Statistical analysis**

Comparisons between the frequent and control donor group with respect to the probability of observing at least one mutation from any gene at VAF threshold 0.5% or 2%, or of at least one *DNMT3A*, *TET2*, *DNMT3A* or *TET2*, Epigenetic or Non-epigenetic gene modifier mutation at VAF threshold 0.5%, were obtained as odds-ratios (OR) based on binomial generalized linear model fits. A quasipoisson generalized linear model accounting for potential over-dispersion was fitted instead to test for the difference in expected number of mutations between the two groups. VAF scores associated with each mutation were compared after log-transformation. Differences in expected log-VAF scores between the two donor groups were tested by fitting linear mixed models, including random intercept terms for donor and gene (where relevant) grouping. All group effects were estimated while controlling for donor age and sequencing depth.

The longitudinal analysis focused on expected VAF score log-fold changes between time-points 1 and 2, which was adopted as response variable. Robust linear modeling was used to account for potential outlier values. The donor group (taking as reference the control donor group), gene (taking as reference *DNMT3A*), donor age at time-point 1, number of days as well as the number of donations between the two donation time-points, and difference in sequencing depth between the two donation time-points, were included as explanatory variables and their effect was therefore estimated mutually adjusted for any other included variable. The VAF log-fold change of donor 444 was imputed from the actual read counts.

CADD scores, fitness scores, stability scores and site mutation rate values were also compared via robust linear modeling; stability scores and site mutation rate values were log-transformed to improve model fitting stability. The models controlled for donor age and included random intercept terms for donor and gene (where relevant) grouping.

For patients having at least one *DNMT3A* mutation, a McNemar exact test was performed to test for the difference in the proportions of presence of a second mutation from the *DNMT3A/TET2* group *vs*. the group of all other epigenetic modifier genes.

All continuous explanatory variables were included after standardization to enhance interpretability and model fitting stability. Analyses were run in R software, v 4.1.1. Linear mixed model fitting was performed via maximum likelihood with the lme4 R package[11] and p-values obtained via the lmerTest R package (Satterthwaite approximation)[12]. The McNemar exact test was calculated with the exact2x2 R package[13]. Robust linear mixed models were fitted with the robustlmm R package[14], and p-values obtained again via the Satterthwaite approximation. Finally, robust modeling without random effects was performed with robustbase R package[15].

Statistical methods used for analysis of in vitro HSPC culture results are outlined in the figure legend. Sample size was not predetermined. Data are presented as means with standard deviation (SD) to indicate the variation within each experiment. For each biological donor a paired t-test was used to compare the percentage of the *DNMT3A*-mutant clones between different conditions.
